# Patient-to-patient phenotype matching enables rare disease gene prioritization beyond curated gene–phenotype associations

**DOI:** 10.64898/2026.01.15.26344236

**Authors:** Isabelle B. Cooperstein, Alistair Ward, Shilpa N. Kobren, Emerson Lebleu, Barry Moore, Rebecca C. Spillmann, Vandana Shashi, Undiagnosed Diseases Network, Gabor T. Marth

**Affiliations:** Department of Human Genetics, University of Utah, Salt Lake City, UT, USA; Frameshift Labs Inc, Cambridge, MA; Department of Biomedical Informatics, Harvard Medical School, Boston, MA; Department of Pediatrics, Division of Medical Genetics, Duke University School of Medicine, Durham, NC

## Abstract

Molecular diagnosis of rare diseases is limited by incomplete knowledge of gene-disease associations, leaving patients with atypical presentations or emerging disorders undiagnosed. We introduce a phenotype-first diagnostic framework that uses previously diagnosed patients as empirical diagnostic evidence, shifting candidate gene prioritization from disease models and predefined gene-phenotype associations to direct patient-to-patient comparison. Implemented as SimPheny, this framework compares an undiagnosed patient with a reference cohort of diagnosed patients and uses the molecular diagnoses of phenotypically similar individuals to prioritize candidate genes, allowing patient-derived clinical evidence to inform diagnosis before association data is incorporated into curated resources. In solved Undiagnosed Diseases Network (UDN) cases, SimPheny outperformed existing prioritization approaches, with the greatest advantage for genes with limited or absent phenotype associations. Clinical review of high-confidence matches in previously unsolved UDN cases confirmed nearly half as diagnostic. These findings establish patient-to-patient matching as a scalable diagnostic strategy for phenotype-driven rare disease diagnostics.

## Introduction

Rare monogenic diseases collectively affect more than 400 million people worldwide, representing a substantial public health burden [1]. Molecular diagnosis of these conditions is essential for clinical management and advancing our understanding of disease mechanisms. Still, causal genes or variants have been identified for fewer than half of the approximately 10,000 recognized rare diseases [2, 3]. Even when disease-associated genes are known, phenotypic heterogeneity and the rarity of individual conditions can complicate recognition of the correct diagnosis [4]. Most patients with a suspected rare genetic disease remain undiagnosed after standard sequencing tests, and many endure prolonged diagnostic odysseys marked by uncertainty and repeated misdiagnoses [5, 6].

These patients typically harbor dozens to hundreds of rare, potentially deleterious variants, of which only one or a small number are truly causative of disease [7]. Variant deleteriousness alone is insufficient to resolve gene-level causality; establishing a diagnosis requires evidence linking a disrupted gene to the patient’s clinical presentation [8]. For well-characterized disease genes with typical presentations, such evidence is generally available, but the task remains challenging for genes that are newly implicated, incompletely characterized, or associated with atypical presentations where the relationship between gene disruption and phenotype is poorly defined.

Phenotypic evidence is therefore critical to variant interpretation, yet its inconsistent application contributes to divergent classifications and reduced diagnostic accuracy [9–12]. The Human Phenotype Ontology (HPO) was developed to standardize phenotypic representation in a structured hierarchy, enabling computational comparisons of phenotypes across patients and diseases [13, 14]. HPO-based annotation is now foundational to rare disease resources including Orphanet [15, 16] and OMIM [17], and has enabled phenotype-aware variant prioritization tools such as Exomiser and AI-MARRVEL, which integrate patient HPO terms with variant pathogenicity predictions, allele frequency, and inheritance patterns to rank candidate genes [18–21]. These approaches have meaningfully improved diagnostic yield, but their effectiveness is bounded by the completeness of existing gene-phenotype association knowledge, leaving many cases inadequately served [22].

Phenotype-based patient matching represents a complementary diagnostic strategy, but existing approaches match a patient’s HPO terms against curated disease models derived from resources such as OMIM or Orphanet [21, 23, 24]. Although these disease models are invaluable references, they represent idealized summaries that rarely capture the full phenotypic variability observed across real patients. Patients with atypical presentations of known conditions may show only weak similarity to the corresponding disease model, risking missed diagnoses [25]. Moreover, patients with novel or insufficiently characterized diseases are not supported by these methods at all, as the relevant disease models do not yet exist.

These limitations motivate a diagnostic framework that, instead of gene-disease associations, uses diagnosed patients as empirical reference models. Rather than comparing a query patient to a curated disease description, a patient-to-patient matching framework identifies previously diagnosed individuals with similar phenotypes and evaluates whether their diagnostic genes overlap with the query patient’s candidate gene list, providing case-level evidence before associations are formally captured in clinical knowledge bases [26, 27]. This approach is grounded in the principle that phenotypic similarity between patients reflects shared genetic etiology. Patient matchmaking platforms such as MatchMaker Exchange have demonstrated the value of connecting patients with similar presentations across institutions to support gene discovery and diagnoses [26], but these efforts have largely functioned as collaborative research tools for identifying novel gene-disease relationships rather than systematic components of routine candidate gene prioritization. Thus, integrating patient-to-patient similarity into the diagnostic pipeline represents a key unmet need.

Here, we present SimPheny, a phenotype-first framework for candidate gene prioritization based on patient-to-patient phenotypic similarity. SimPheny queries a reference cohort of diagnosed patients using HPO-encoded phenotypes and scores and ranks genes in the query patient’s candidate gene list that overlap with the diagnostic genes of the reference patients. We evaluate this framework using deeply phenotyped participants from the Undiagnosed Diseases Network (UDN), benchmark its performance against established prioritization approaches, and demonstrate its utility for identifying clinically actionable diagnostic candidates in previously unsolved UDN cases.

## Results

### A patient-to-patient diagnostic framework for candidate gene prioritization

We developed SimPheny as an implementation of a patient-to-patient phenotype matching framework for diagnostic gene prioritization, using previously diagnosed patients as empirical reference models. SimPheny accepts two inputs for each undiagnosed “query” patient: (i) a set of Human Phenotype Ontology (HPO) terms describing the patient’s clinical features and (ii) a candidate list of genes harboring variants with predicted functional impact, derived from sequencing-based variant prioritization (**Fig. 1**; <u>Methods</u>). Using these inputs, SimPheny computes pairwise phenotypic similarity between the query patient and all individuals in a diagnosed reference cohort using PhenoSim_Jaccard_, a semantic similarity metric that incorporates HPO structure and term specificity (<u>Methods</u>).

**Figure 1:**
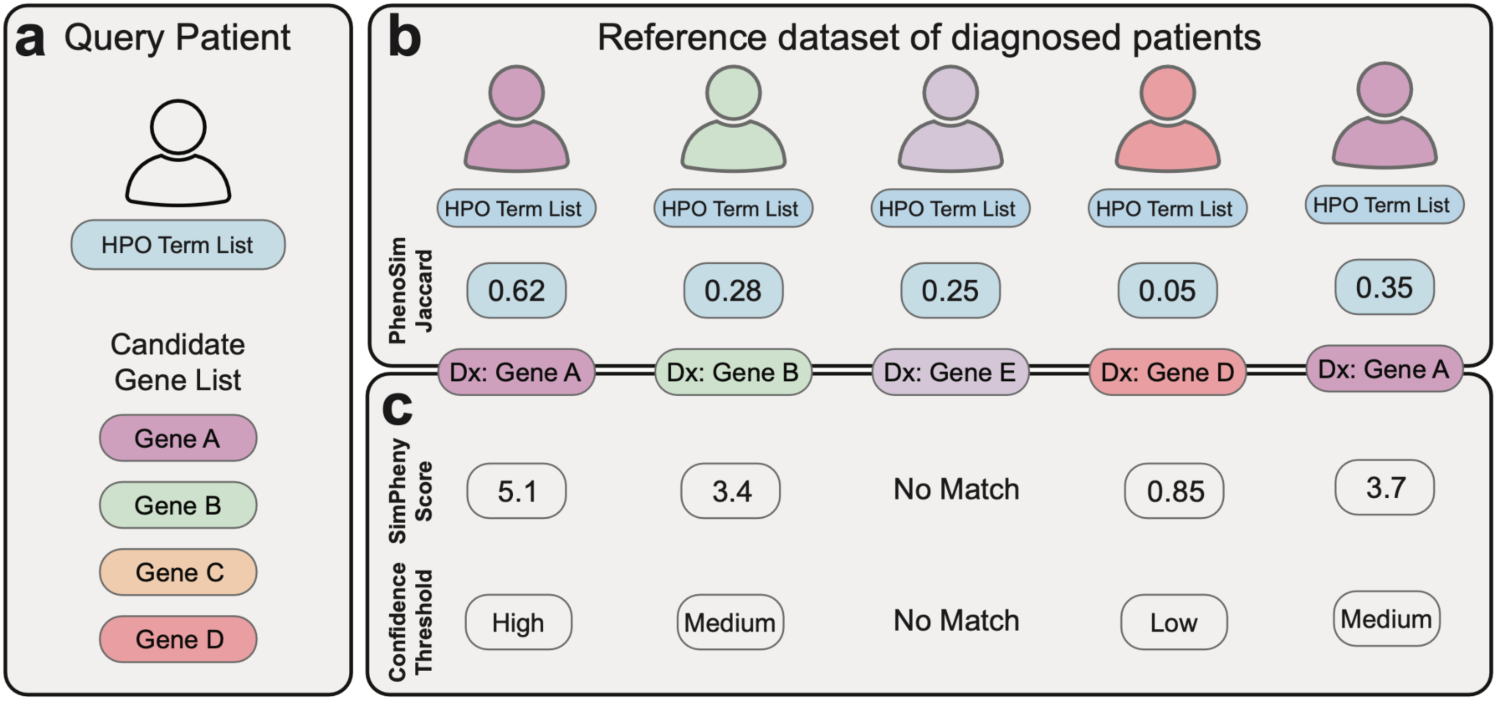
Patient-to-patient phenotype matching framework for candidate gene prioritization. **a)** Query patient inputs: HPO terms describing the clinical presentation and a candidate gene list from sequencing analysis. **b)** Pairwise phenotypic similarity is calculated between the query patient and all individuals in a diagnosed reference dataset using PhenoSim_Jaccard_. **c)** Overlaps between the query patient’s candidate genes and diagnostic genes of reference patients define SimPheny Matches, each assigned a SimPheny Score integrating phenotypic similarity and gene-level specificity. Matches are categorized into high-, medium-, and low-confidence tiers for clinical interpretation. Reference patients whose diagnostic genes do not overlap the candidate gene list produce no match. Dx = diagnosis.

SimPheny then identifies reference patients whose diagnostic genes overlap the query patient’s candidate gene list, defining each overlap as a *SimPheny Match*. For each match, a *SimPheny Score* is computed by integrating the statistical significance of the pairwise phenotypic similarity with the specificity of the shared gene (<u>Methods</u>). Matches are assigned to high-, medium-, or low-confidence tiers based on benchmarking-derived thresholds to support clinical interpretation.

### The UDN provides a phenotypically diverse reference cohort for patient-to-patient matching

To benchmark and apply SimPheny, we used data from the Undiagnosed Diseases Network (UDN), a national research study established to improve diagnosis and discovery for rare and undiagnosed conditions [28]. The study cohort comprised 2,214 UDN participants with HPO-encoded phenotypes, including 767 diagnosed probands or affected relatives and 1,447 undiagnosed probands (**Supplementary Fig. 1**; <u>Methods</u>). Participants were annotated with a median of 16 HPO terms describing their clinical presentations (**Supplementary Fig. 2**). The cohort spanned a broad range of genetic disorders affecting multiple organ systems, reflecting the phenotypic heterogeneity characteristic of rare disease populations and providing a robust setting for evaluating patient-to-patient matching.

### The Jaccard similarity metric captures structured phenotypic relationships among disease models

A central requirement for patient-to-patient diagnostic matching is a phenotypic similarity metric that recognizes not only exact phenotype matches, but also biologically meaningful relationships between different but related HPO terms. SimPheny uses *PhenoSim_Jaccard_*, a semantic similarity metric that extends the standard Jaccard index by incorporating ontology structure and term information content, such that similarity between two HPO terms reflects the specificity of their shared ancestors relative to all ancestors across both terms (<u>Methods</u>; **Supplementary Fig. 3**).

Term-to-term similarities are aggregated into set-level scores using *funSimAvg* [29], which averages best-matching terms between two HPO sets in both directions, yielding a score between zero and one, one indicating an exact match. This structure rewards shared ancestry between terms, particularly when shared ancestors are specific and informative.

To evaluate whether *PhenoSim_Jaccard_* captures clinically meaningful phenotypic structure, we computed pairwise similarities across 4,120 Orphanet disease models [16] (**Supplementary Fig. 4**). Visualization by the commonly used t-SNE nonlinear dimensionality reduction method revealed organization by top-level HPO categories, with distinct groupings such as eye and cardiovascular disorders emerging (**Supplementary Fig. 5a–b**; <u>Methods</u>). Consistent with this structure, disease pairs within the same top-level HPO category exhibited significantly higher *PhenoSim_Jaccard_* scores than pairs across different categories (**Fig. 2a**). Together, these results demonstrate that *PhenoSim_Jaccard_* preserves the higher-order phenotypic structure encoded in the HPO, supporting its use as a similarity metric for patient-level phenotypic matching.

**Figure 2:**
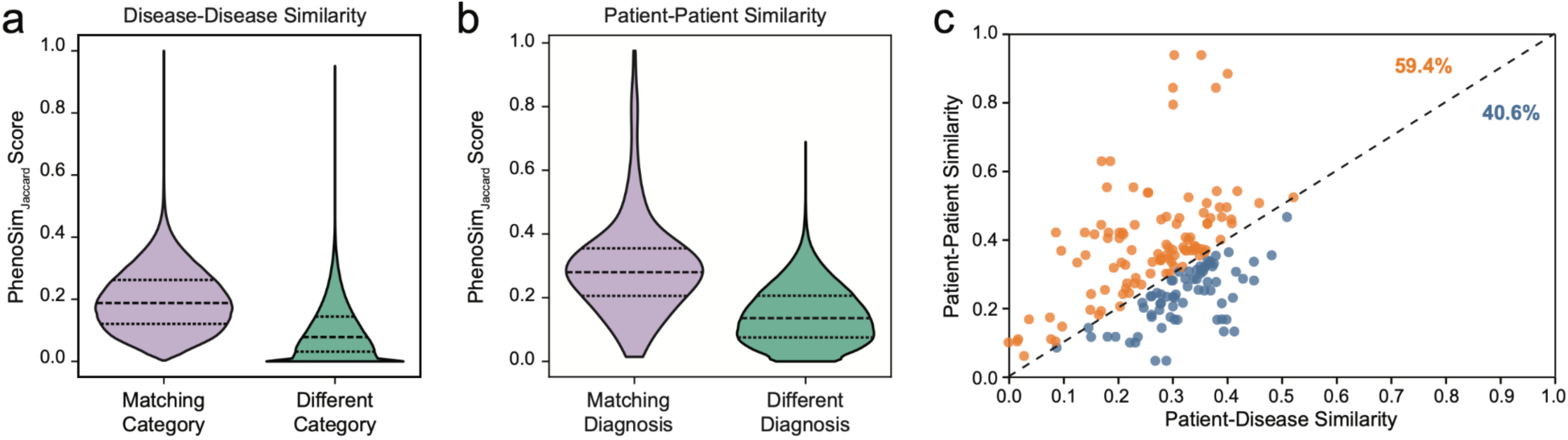
PhenoSim_Jaccard_ measures phenotypic similarity in disease models and patients. a) Pairwise PhenoSim_Jaccard_ scores between 4,120 Orphanet disease models, stratified by whether pairs share the same top-level HPO disease category (*Matching Category*) or belong to different categories (*Different Category*). b) Pairwise patient-to-patient PhenoSim_Jaccard_ scores among 767 diagnosed UDN participants, stratified by whether pairs share the same genetic diagnosis (*Matching Diagnosis*) or different genetic diagnoses (*Different Diagnoses*). c) Subset of 175 diagnosed UDN participants for whom (i) at least one additional UDN participant shared the same diagnostic gene and (ii) an Orphanet disease model was available for that gene. For each participant, the highest-scoring patient-to-patient match (y-axis) is plotted against the highest-scoring patient-to-disease match with the corresponding Orphanet model (x-axis). Points are colored by whether the patient-to-patient score exceeded the patient-to-disease score (orange; above diagonal) or not (blue, below diagonal). The dashed line marks equal similarity (x = y). Patient-to-patient similarity exceeded patient-to-disease similarity for the majority of participants (59.4%; mean signed difference = 0.043, median = 0.028; Wilcoxon signed-rank test, p = 0.0059).

### Patient-to-patient phenotypic similarity enriches for shared genetic diagnoses

Curated disease resources represent each condition as a single HPO term set. Although these disease models provide valuable diagnostic references, they cannot fully capture the phenotypic variability observed across individuals sharing the same molecular diagnosis, and no such model exists for patients whose underlying disease has not yet been formally described. We therefore evaluated whether patient-to-patient phenotypic similarity could distinguish patients with shared genetic diagnoses within the UDN cohort and whether diagnosed patients provided more similar phenotypic references than curated disease models.

Among the 767 diagnosed UDN participants in our reference cohort, pairwise *PhenoSim_Jaccard_* scores were higher for participants sharing the same genetic diagnosis than for participants with different diagnoses (**Fig. 2b**), demonstrating that patient-to-patient phenotypic similarity is enriched among individuals with shared genetic diagnoses. To directly compare patient-to-patient and patient-to-disease similarity, we identified 175 diagnosed UDN participants for whom both an Orphanet disease model corresponding to their diagnostic gene and at least one additional UDN participant diagnosed in the same gene were available. For each participant, we compared the highest *PhenoSim_Jaccard_* score among same-gene patient matches with the *PhenoSim_Jaccard_* score against the corresponding Orphanet disease model. Patient-to-patient similarity exceeded patient-to-disease similarity for 59.4% of participants (Wilcoxon signed-rank test, p = 0.0059; **Fig. 2c**).

These findings demonstrate that diagnosed patients can provide phenotype representations comparable to, and in many cases more similar than, curated disease models. Patient-level reference data therefore provide empirical representations of phenotypic variation that may not be captured by a single curated disease description.

### Patient-to-patient matching effectively prioritizes diagnostic genes in previously solved UDN cases

We next benchmarked SimPheny using previously solved UDN cases to evaluate its ability to prioritize diagnostic genes from patient-specific candidate gene lists. For benchmarking, we focused on diagnosed UDN probands with jointly called family-level variant data available (**Supplementary Fig. 1**; <u>Methods</u>). For each proband, we generated an unranked candidate gene list from family-level variant data using Exomiser [18], consistent with prior studies that use Exomiser-derived candidate gene lists as input to downstream diagnostic prioritization methods [30, 31]. Exomiser filters variants based on inheritance patterns and population frequency and annotates them using predicted pathogenicity and phenotype association scores. We retained genes for which either the variant-level or phenotype-level score exceeded a minimum threshold (<u>Methods</u>). This approach retains genes with strong variant evidence despite limited or absent phenotypic support, allowing downstream methods such as SimPheny to evaluate candidate genes that association-based prioritization approaches would otherwise rank poorly.

Benchmarking required that each proband’s known diagnostic gene be present in the candidate gene list, as SimPheny cannot rank genes absent from the input. Of the diagnosed probands with available variant data, 404 met this criterion and comprised the final benchmarking cohort. Excluded probands largely had diagnoses attributable to structural variants, which were not represented in the SNV/indel VCFs, or noncoding variants, which were not evaluated by the Exomiser-based workflow used to generate candidate gene lists for this analysis (**Supplementary Fig. 6**; <u>Methods</u>) [32]. As SimPheny requires a patient-specific candidate gene list for benchmarking, these cases could not be evaluated. Nine probands contributed more than one molecular diagnosis, yielding 415 diagnostic gene instances for evaluation. Because the same gene could appear in the candidate lists of multiple probands, gene instances rather than unique genes were counted. Candidate gene lists additionally contained 29,543 non-diagnostic gene instances across the cohort, with a median of 41 candidate genes per proband (**Supplementary Fig. 7a**).

We applied SimPheny to each benchmarking proband using their HPO terms and candidate gene list, comparing each case against the 767 diagnosed UDN reference participants while excluding self-matches (**Fig. 1; Supplementary Fig. 1**). A *SimPheny Match* was identified when a candidate gene overlapped the diagnostic gene of a reference participant. For each match, a *SimPheny Score* was calculated by integrating the statistical significance of the phenotypic similarity with the cohort-wide frequency of the shared gene (<u>Methods</u>). Candidate matches were prioritized by decreasing *SimPheny Score*, while genes without any matching diagnosed reference patients received no score and therefore remained unprioritized. Matches involving the proband’s known diagnostic gene were classified as true positives (TPs), whereas all other matches were classified as false positives (FPs).

SimPheny identified 3,442 matches across the benchmarking probands (**Fig. 3c**). Of the 415 diagnostic gene instances, 156 (37.6%) had at least one additional diagnosed case with the same true diagnostic gene in the UDN reference cohort and were therefore recoverable through patient-to-patient matching. Because only genes represented in the diagnosed reference cohort receive a *SimPheny Score*, the breadth of the reference cohort is a primary determinant of diagnostic coverage. Benchmarking probands had a median of seven matches across three unique genes (**Supplementary Fig. 8**), reducing the median candidate list from 41 input genes to three phenotype-informed diagnostic candidates (**Supplementary Fig. 7a**).

**Figure 3:**
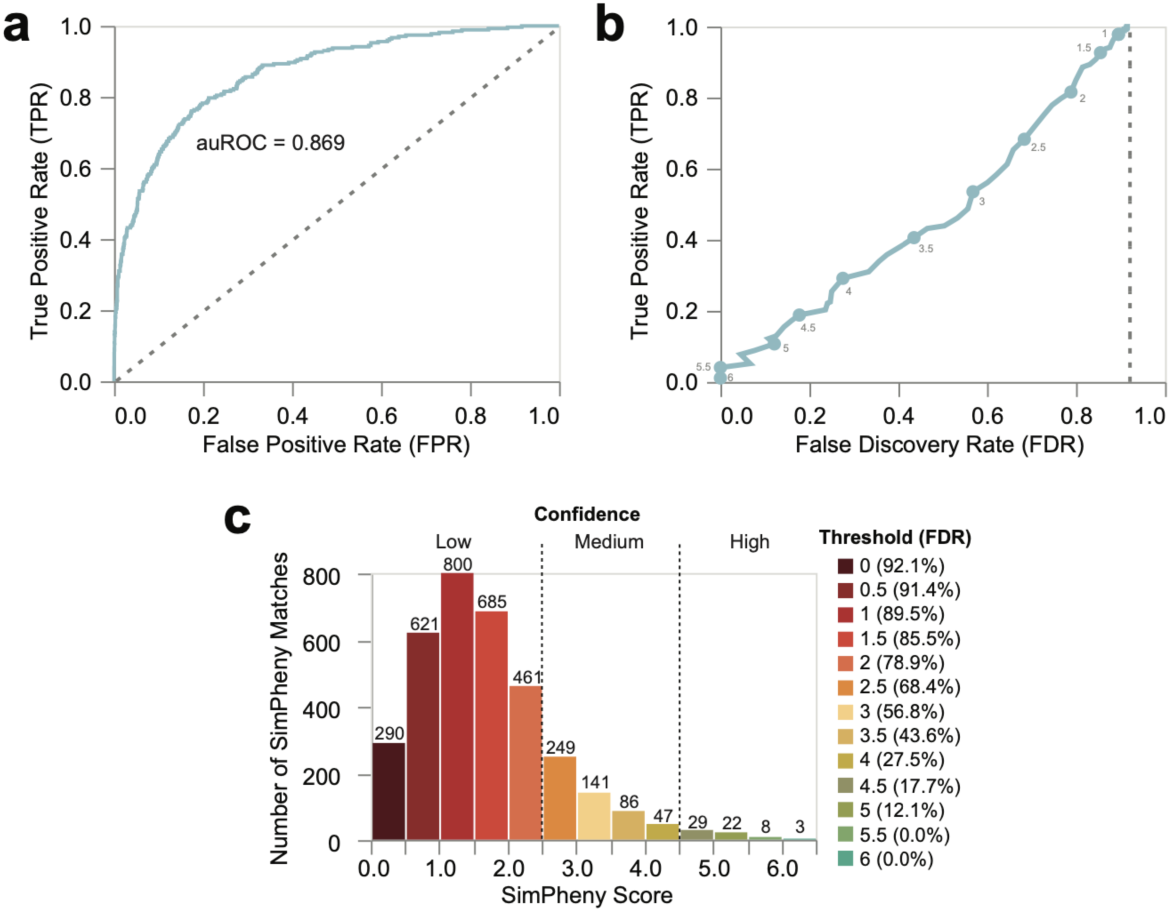
Establishing confidence thresholds for *SimPheny Matches* in the benchmarking cohort. **a)** Receiver Operating Characteristic (ROC) curve evaluating *SimPheny Scores*’ ability to distinguish true diagnostic matches from false positives in the UDN benchmarking cohort. The dashed line represents a random classifier. **b)** False Discovery Rate (FDR) curve. Points correspond to *SimPheny Score* thresholds in 0.5 increments, ranging from 1 (top right) to 6.0 (bottom left). The dashed line represents the random expectation, where 92.1% of matches would be predicted as false due to the class imbalance. **c)** Distribution of *SimPheny Scores* across all 3,442 matches in the benchmarking cohort, binned in 0.5 increments. Scores range from 0.06 to 6.36. Higher scores reflect stronger joint evidence that the observed patient-to-patient phenotypic similarity and gene overlap are unlikely to occur by chance. Dashed lines mark the three confidence tiers defined by FDR analysis: high (≥ 4.5), medium (2.5-4.5), and low (< 2.5). Numbers above bars indicate the number of matches within each bin. Median *SimPheny Score* = 1.51.

We next evaluated whether *SimPheny Scores* distinguished diagnostic (TP) from non-diagnostic (FP) matches. Receiver operating characteristic (ROC) analysis yielded an area under the curve (AUC) of 0.869 (**Fig. 3a**), indicating that *SimPheny Scores* effectively discriminated TP from FP matches. As fewer than 10% of the returned matches were TPs, we used false discovery rate (FDR) as the primary performance metric in this highly imbalanced setting (<u>Methods</u>). At a permissive threshold of *SimPheny Score* ≥ 1.5, SimPheny recovered 92.6% of TPs with an FDR of 85.5%, maximizing sensitivity at the cost of precision (**Fig. 3b**; **Table 1**; **Supplementary Table 1**). At the stringent threshold of *SimPheny Score* ≥ 4.5, SimPheny recovered 18.8% of TPs while reducing the FDR to 17.7%, meaning that approximately four of every five reported matches involved the diagnostic gene. We therefore defined three confidence tiers: high-confidence (*SimPheny Score* ≥ 4.5), medium-confidence (2.5 ≤ *SimPheny Score* < 4.5), and low-confidence (*SimPheny Score* < 2.5) (**Table 1**; **Supplementary Table 1**), allowing users to balance sensitivity and precision according to clinical or research objectives.

**Table 1:**
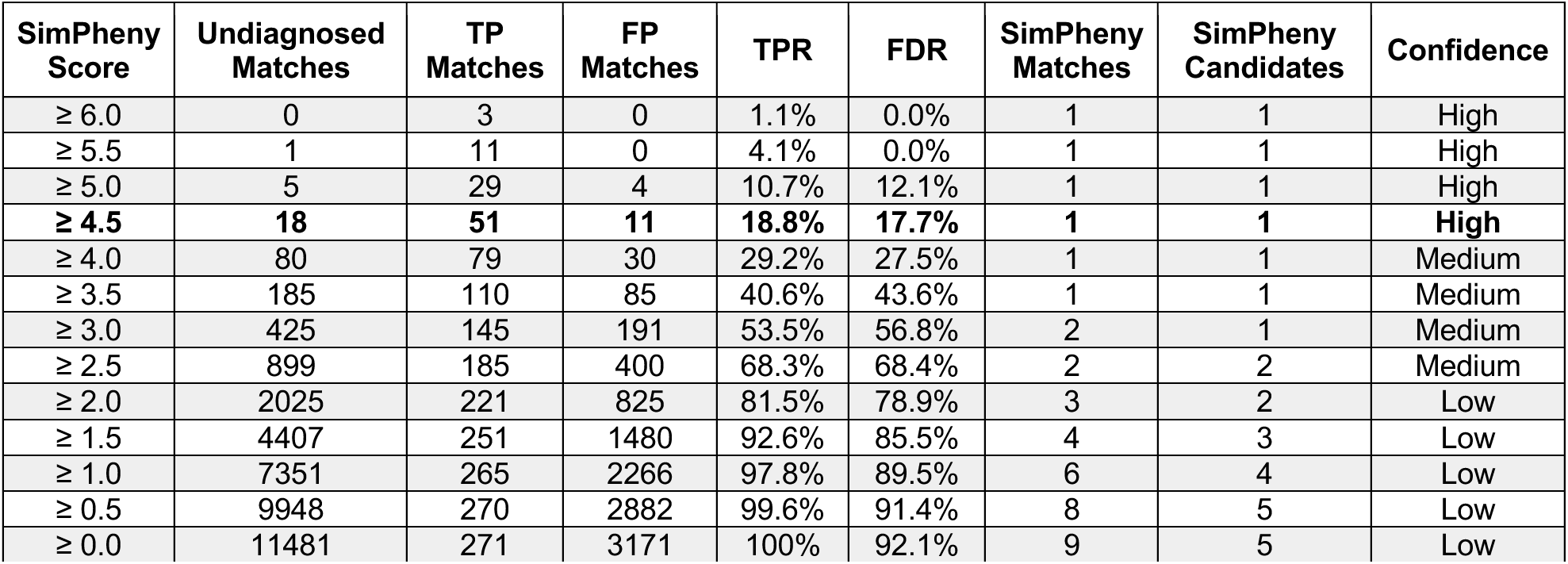
Summary statistics for SimPheny Matches across varying SimPheny Score thresholds in benchmarking and discovery cohorts. SimPheny was run on 1,851 UDN probands – 404 diagnosed individuals in the benchmarking cohort and 1,447 undiagnosed individuals in the discovery cohort – querying against a reference dataset of 767 diagnosed UDN participants (**Supplementary Fig. 1**). Each row summarizes results at a given *SimPheny Score* threshold in the leftmost column. True positive (TP) and false positive (FP) matches were determined in the benchmarking cohort of 404 diagnosed probands with known diagnostic genes. “Undiagnosed Matches” refers to matches returned from the 1,447 undiagnosed probands. TPR (true positive rate) and FDR (false discovery rate) were calculated from the benchmarking data (**Fig. 3**). “SimPheny Matches” and “SimPheny Candidates” indicate the average number of matches or candidate genes per proband (of 1,851) among those with at least one match at each score threshold. Confidence tiers were derived from benchmarking performance.

| SimPheny Score | Undiagnosed Matches | TP Matches | FP Matches | TPR | FDR | SimPheny Matches | SimPheny Candidates | Confidence |
| --- | --- | --- | --- | --- | --- | --- | --- | --- |
| ≥ 6.0 | 0 | 3 | 0 | 1.1% | 0.0% | 1 | 1 | High |
| ≥ 5.5 | 1 | 11 | 0 | 4.1% | 0.0% | 1 | 1 | High |
| ≥ 5.0 | 5 | 29 | 4 | 10.7% | 12.1% | 1 | 1 | High |
| **≥ 4.5** | **18** | **51** | **11** | **18.8%** | **17.7%** | **1** | **1** | **High** |
| ≥ 4.0 | 80 | 79 | 30 | 29.2% | 27.5% | 1 | 1 | Medium |
| ≥ 3.5 | 185 | 110 | 85 | 40.6% | 43.6% | 1 | 1 | Medium |
| ≥ 3.0 | 425 | 145 | 191 | 53.5% | 56.8% | 2 | 1 | Medium |
| ≥ 2.5 | 899 | 185 | 400 | 68.3% | 68.4% | 2 | 2 | Medium |
| ≥ 2.0 | 2025 | 221 | 825 | 81.5% | 78.9% | 3 | 2 | Low |
| ≥ 1.5 | 4407 | 251 | 1480 | 92.6% | 85.5% | 4 | 3 | Low |
| ≥ 1.0 | 7351 | 265 | 2266 | 97.8% | 89.5% | 6 | 4 | Low |
| ≥ 0.5 | 9948 | 270 | 2882 | 99.6% | 91.4% | 8 | 5 | Low |
| ≥ 0.0 | 11481 | 271 | 3171 | 100% | 92.1% | 9 | 5 | Low |

### Patient-derived phenotypic evidence improves candidate gene ranking beyond curated associations

As existing prioritization tools generally rely on documented relationships between genes, phenotypes, and diseases from resources such as OMIM and Orphanet, we hypothesized that SimPheny’s use of patient-derived phenotypic evidence would more effectively prioritize genes that are newly described, sparsely annotated, or present atypically.

Accordingly, we compared SimPheny with three widely used tools representing distinct prioritization strategies: Phrank [33], which scores phenotypic similarity between patient HPO terms and OMIM/Orphanet gene–phenotype associations; AI-MARRVEL [20], a machine learning framework that integrates OMIM-based gene–phenotype similarity with variant-level and literature-derived features; and Exomiser [18], which combines variant pathogenicity predictions with phenotypic similarity derived from OMIM/Orphanet associations. Exomiser was evaluated using both its default parameters and our previously optimized UDN-specific settings, which restrict associations to human data [32]. Exomiser was used upstream solely as a filtering step to generate reasonable candidate gene lists for benchmarking; here, we evaluated its gene rankings as an independent comparison method.

As SimPheny relies on the phenotypic similarity between patients, we first evaluated whether *PhenoSim_Jaccard_* more effectively captures diagnostic similarity than Phrank’s similarity metric. *PhenoSim_Jaccard_* assigned higher phenotypic similarity ranks to patients sharing the same diagnostic gene and produced greater separation between matching and non-matching diagnosis pairs than Phrank-based similarity (**Supplementary Fig. 9a–b**). We next compared Phrank gene scores with *SimPheny Scores* across the 156 diagnostic gene instances for which patient-to-patient matching was possible in our benchmarking cohort. SimPheny demonstrated greater discrimination than Phrank by ROC analysis (auROC = 0.863 versus 0.801) and maintained lower FDR across comparable TPRs (**Supplementary Fig. 9c–d**), supporting the use of direct patient-derived phenotypic evidence over curated gene associations for candidate gene scoring.

To compare overall gene prioritization performance, candidate genes were ranked using a *SimPheny Gene Score* defined as the average of the two highest *SimPheny Scores* for each gene across all reference matches (<u>Methods</u>). Across the 156 diagnostic gene instances with at least one corresponding diagnosed reference case, SimPheny ranked 116 (74.4%) as the top candidate and 150 (96.2%) within the top five, compared with 112 (71.8%) and 131 (84.0%) by AI-MARRVEL (**Fig. 4a**; **Supplementary Table 2**). SimPheny also outperformed both Exomiser configurations and never ranked a diagnostic gene below the top 15. In contrast, the comparison methods ranked some diagnostic genes beyond the top 50 or failed to recover them at all. Including multispecies phenotype associations under Exomiser’s default settings did not improve ranking relative to the human-only optimized settings, suggesting that cross-species phenotype data contributed little additional diagnostic signal.

**Figure 4:**
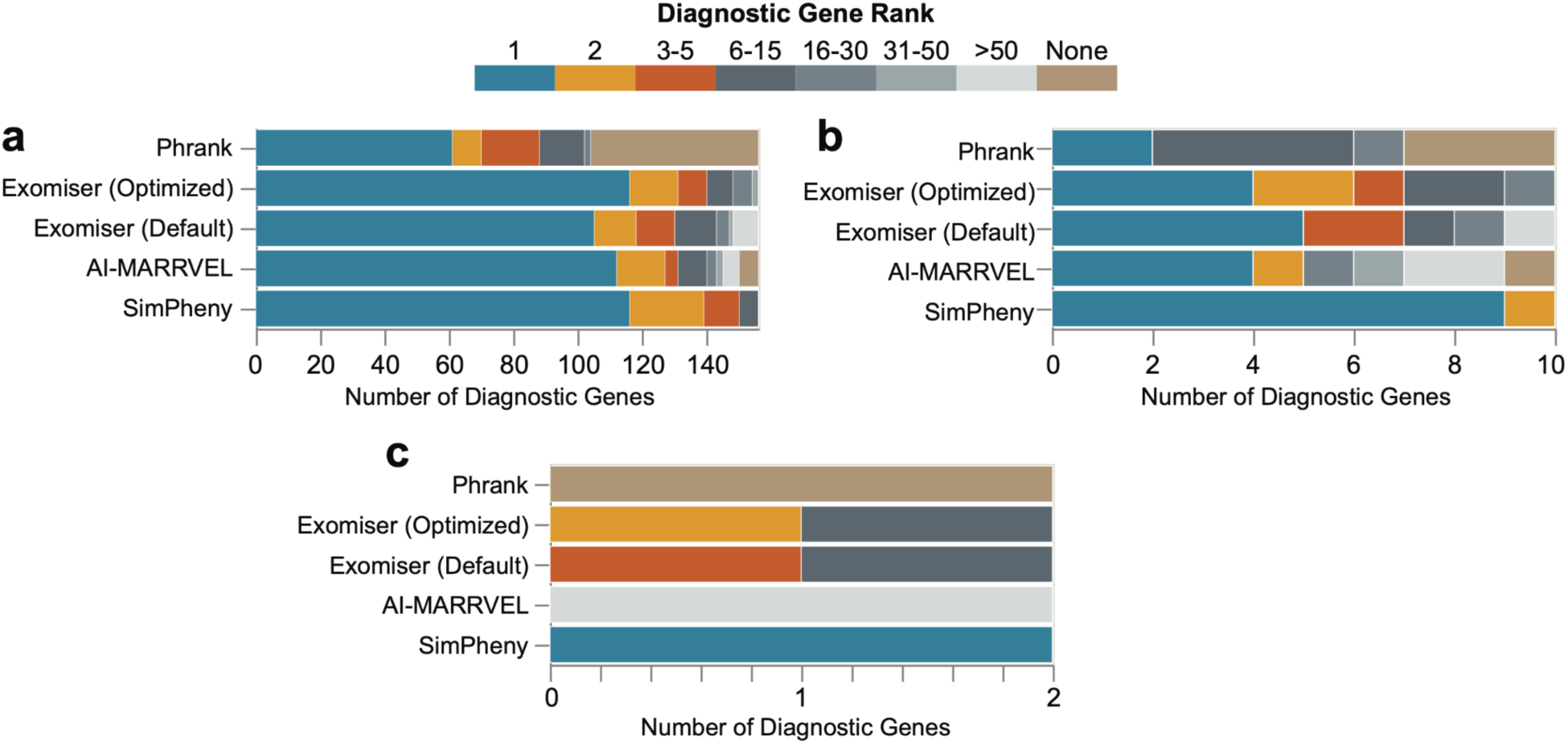
SimPheny outperforms state-of-the-art candidate gene prioritization methods. Performance of SimPheny versus Phrank, Exomiser (UDN-optimized and default parameters; see Methods), and AI-MARRVEL. Performance was measured as the rank assigned to diagnostic genes by each tool. **a)** Diagnostic genes with at least one true positive match in the UDN reference dataset (n=156) **b)** Diagnostic genes whose associated disease descriptions in OMIM or Orphanet had no direct phenotypic overlap with the proband’s HPO terms (n=10). **c)** Diagnostic genes with *no* documented HPO associations in OMIM or Orphanet (n=2).

The benefit of patient-derived phenotypic evidence was most evident for genes with limited curated phenotypic associations. Among ten diagnostic gene instances whose associated OMIM or Orphanet disease descriptions shared no HPO terms with the proband, SimPheny ranked all ten among the top two candidates, whereas other methods ranked some beyond the top 50 or failed to rank them entirely (**Fig. 4b**). For two probands whose diagnostic genes had *no* associated HPO terms in OMIM or Orphanet, SimPheny ranked both as the top candidate, whereas AI-MARRVEL ranked them below 50th, and Phrank could not score them as it can only evaluate genes annotated in OMIM or Orphanet (**Fig. 4c**).

### Patient-to-patient matching identifies diagnoses in previously unsolved UDN probands

After benchmarking the framework in solved cases, we applied SimPheny to the 1,447 undiagnosed probands in the discovery cohort, querying against the 767 diagnosed participants in the UDN reference cohort (**Fig. 1**; **Supplementary Fig. 1**). SimPheny identified 11,481 candidate gene matches across the discovery cohort, returning a median of six matches across three unique candidate genes per proband – substantially reducing the candidate space from the median of 42 genes in the original candidate lists (**Fig. 5a–b**; **Supplementary Fig. 7b**). Of the 1,447 probands, 1,250 (86.4%) yielded at least one gene match in the UDN reference cohort.

**Figure 5:**
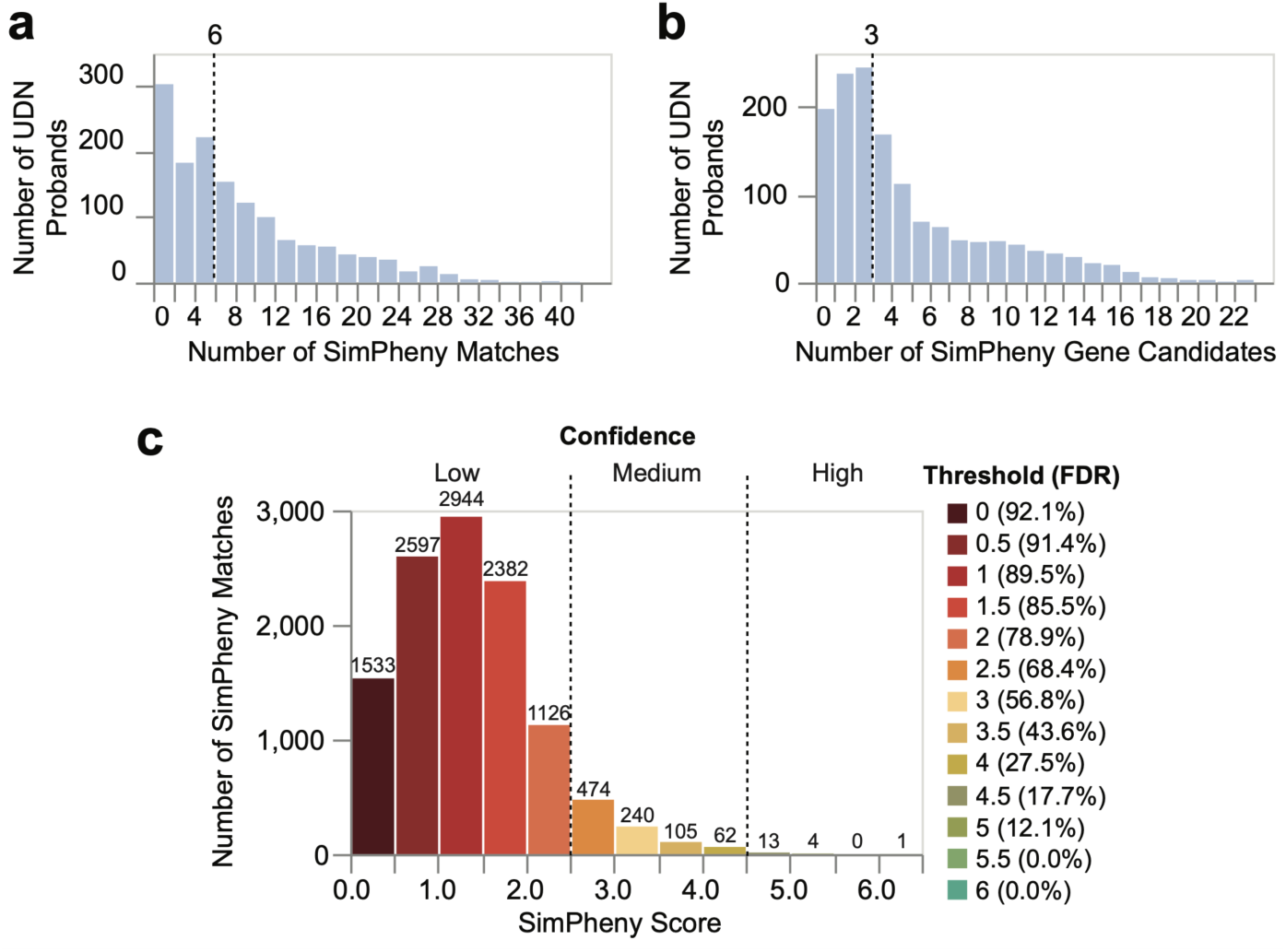
SimPheny prioritizes candidate genes for diagnostic review in the UDN discovery cohort. **a)** Number of SimPheny Matches returned per proband when querying 1,447 undiagnosed probands against 767 diagnosed UDN reference participants. A single proband may have multiple matches to the same gene if multiple reference cases are diagnosed in that gene. The dashed line indicates the median number of matches per proband. 197 probands (13.6%) had no matches. **b)** Number of unique SimPheny gene candidates returned per proband. The dashed line indicates the median number of candidate genes per proband. **c)** Distribution of SimPheny Scores across 11,481 matches in the discovery cohort, binned in 0.5 increments. Scores range from 0.03 to 6.09. Dashed lines mark confidence tiers defined by FDR benchmarking analyses (Fig. 3; **Supplementary Table 1**): high (≥ 4.5), medium (2.5-4.5), and low (<2.5). Numbers above bars indicate counts per bin. Median SimPheny Score = 1.27. The 18 high-confidence matches correspond to the 17 SimPheny candidates prioritized for diagnostic review.

We prioritized high-confidence matches with *SimPheny Score* ≥ 4.5 for manual review, corresponding to an estimated FDR of 17.7% from the benchmarking cohort (**Table 1**; **Fig. 3**). This threshold yielded 18 high-confidence matches across 17 probands; one proband had two high-confidence matches to the same gene, *RPL13*, which were reviewed as a single candidate.

Each high-confidence candidate underwent manual review, including assessment of phenotypic concordance with the matched reference participant, inheritance patterns, variant pathogenicity predictions, ClinVar assertions, and other supporting clinical evidence (**Table 2**; **Supplementary Table 3**).

**Table 2:**
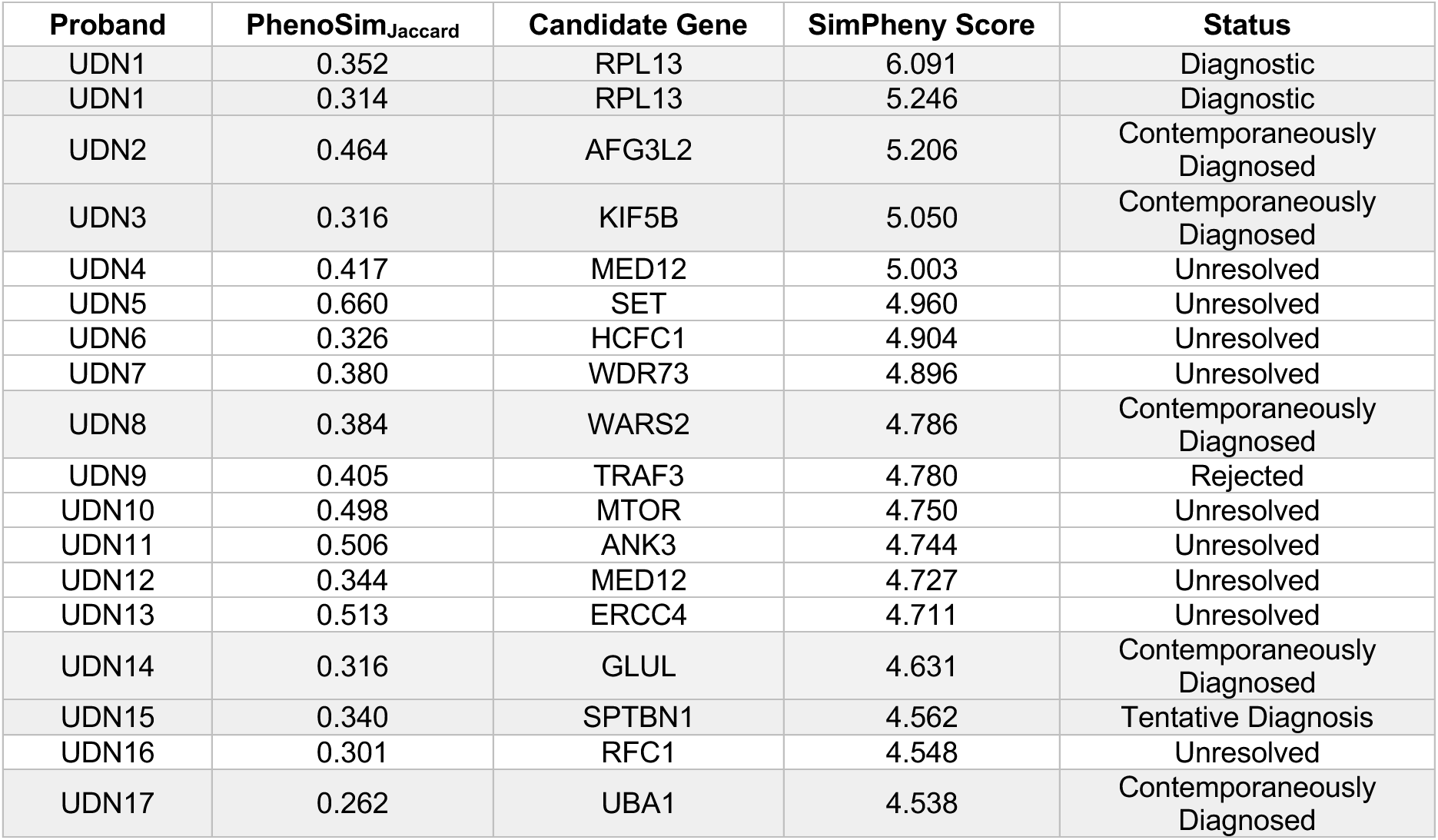
High-confidence candidate gene matches (SimPheny Score ≥ 4.5) in undiagnosed UDN probands. SimPheny was applied to 1,447 undiagnosed UDN probands in the discovery cohort, querying against a reference dataset of 767 diagnosed UDN participants (**Supplementary Fig. 1**). Each row represents a high-confidence SimPheny Match, including the phenotypic similarity score (*PhenoSim_Jaccard_*), *SimPheny Score*, and classification status based on manual review. “Diagnostic” refers to candidates newly identified by SimPheny that are now considered diagnostic by the UDN clinical team. “Contemporaneously Diagnosed” matches correspond to diagnoses independently reached by UDN sites but not recorded in the UDN internal database at the time of cohort curation. “Tentative Diagnosis” denotes a match in which the candidate gene was recorded in the UDN internal database but excluded from our diagnosed cohort due to diagnostic uncertainty (Methods). “Rejected” candidates were ruled out by UDN clinical review. “Unresolved” candidates refer to cases where SimPheny candidates lack sufficient evidence to be either rejected or confirmed, although we note that some have benign/likely benign variant classifications in ClinVar. See **Supplementary** Table 3 for extended match details.

| Proband | $PhenoSim_{Jaccard}$ | Candidate Gene | SimPheny Score | Status |
| --- | --- | --- | --- | --- |
| UDN1 | 0.352 | RPL13 | 6.091 | Diagnostic |
| UDN1 | 0.314 | RPL13 | 5.246 | Diagnostic |
| UDN2 | 0.464 | AFG3L2 | 5.206 | Contemporaneously Diagnosed |
| UDN3 | 0.316 | KIF5B | 5.050 | Contemporaneously Diagnosed |
| UDN4 | 0.417 | MED12 | 5.003 | Unresolved |
| UDN5 | 0.660 | SET | 4.960 | Unresolved |
| UDN6 | 0.326 | HCFC1 | 4.904 | Unresolved |
| UDN7 | 0.380 | WDR73 | 4.896 | Unresolved |
| UDN8 | 0.384 | WARS2 | 4.786 | Contemporaneously Diagnosed |
| UDN9 | 0.405 | TRAF3 | 4.780 | Rejected |
| UDN10 | 0.498 | MTOR | 4.750 | Unresolved |
| UDN11 | 0.506 | ANK3 | 4.744 | Unresolved |
| UDN12 | 0.344 | MED12 | 4.727 | Unresolved |
| UDN13 | 0.513 | ERCC4 | 4.711 | Unresolved |
| UDN14 | 0.316 | GLUL | 4.631 | Contemporaneously Diagnosed |
| UDN15 | 0.340 | SPTBN1 | 4.562 | Tentative Diagnosis |
| UDN16 | 0.301 | RFC1 | 4.548 | Unresolved |
| UDN17 | 0.262 | UBA1 | 4.538 | Contemporaneously Diagnosed |

Of the 17 independently reviewed candidate gene matches, 7 (41.2%) were confirmed as diagnostic. Five of seven corresponded to diagnoses independently reached by UDN clinical teams but not yet incorporated into the internal UDN database at the time of cohort curation; these are hereafter referred to as *contemporaneously diagnosed* candidates. One additional match corresponded to the *RPL13* diagnosis described below, which the UDN credited to SimPheny. The final confirmed case was a SimPheny candidate corresponding to a molecular diagnosis previously proposed by the UDN; however, the proband was classified as undiagnosed during cohort curation because this finding was not considered definitive (<u>Methods</u>).

SimPheny ranked *RPL13* as the top candidate in a proband with two high-confidence *RPL13* matches and a third match just below threshold (SimPheny Score = 4.45). This proband and family had been evaluated by the UDN five years earlier with a clinical impression of unclassified spondyloepimetaphyseal dysplasia without an identified genetic cause. Communication with the UDN clinical site and reporting laboratory revealed that the family’s genome sequencing had been analyzed before *RPL13* was first implicated in Spondyloepimetaphyseal dysplasia, Isidor-Toutain type. The same *RPL13* variant identified in this family was reported in the original publication that established the disorder [34] and, later, in an independent study [35], confirming its pathogenicity. SimPheny’s nomination of *RPL13* prompted segregation testing, which confirmed the variant in the proband and four affected relatives, while an unaffected sibling tested negative. This molecular diagnosis was subsequently returned to the family as a certain diagnosis, credited to SimPheny by the UDN.

In another proband with unexplained osteogenesis imperfecta, Exomiser deprioritized *KIF5B*, assigning it a phenotype score of zero, consistent with the newly described disease association not yet being represented in the underlying phenotype resources used by Exomiser (**Supplementary Table 3**). SimPheny identified *KIF5B* as the top candidate by matching the proband to a diagnosed reference participant with KIF5B-related osteogenesis imperfecta. This case was contemporaneously diagnosed by the UDN clinical team, illustrating how patient-to-patient matching can identify newly recognized disease genes before curated gene-phenotype resources fully capture emerging evidence.

Of the ten remaining high-confidence matches, the overseeing clinical team rejected one, and nine remain unresolved pending further evaluation. Among the nine unresolved candidates, five involve variants classified as benign or likely benign in ClinVar [36] and are therefore likely to be rejected on further review. Two involve conflicting ClinVar interpretations, one is classified as a variant of uncertain significance, and one is not currently represented in ClinVar (**Supplementary Table 3**).

### Increasing the size and gene representation of the reference cohort increases the diagnostic coverage of patient-to-patient matching

The diagnostic power of patient-to-patient matching is inherently constrained by the composition of the diagnosed reference cohort: a true diagnostic match cannot be identified if no diagnosed reference patient exists for that gene. Therefore, increasing the size of the diagnosed reference cohort – and consequently the number of diagnostic genes represented – should increase the likelihood that an undiagnosed patient finds a relevant match. To quantify this effect, we compared diagnostic coverage using the UDN-only reference cohort of 767 participants and an expanded reference cohort incorporating three additional resources: 6,451 cases from the Phenopacket Store [37], 11,797 ClinVar pseudopatients derived from variant-level submissions (excluding UDN submissions) [36], and 4,576 patients from DECIPHER [38], yielding a combined reference cohort of 23,591 diagnosed cases (**Supplementary Methods**).

Expanding the reference cohort increased the number of represented diagnostic genes from 592 in the UDN-only cohort to 2,939 across all reference datasets (**Supplementary Table 4**). As a result, the number of benchmarking diagnostic gene instances with at least one corresponding diagnosed reference patient increased from 156 (37.6%) to 354 (85.3%) (**Fig. 6a**; **Supplementary Table 4**). Although the larger reference cohort increased both the total number of patient-to-patient matches and the number of SimPheny candidate genes per proband (**Supplementary Fig. 10**), ranking performance remained strong: 313 of the 354 (88.4%) diagnostic gene instances represented by a diagnosed patient in the expanded reference cohort were ranked among the top five SimPheny candidates (**Fig. 6a**).

**Figure 6:**
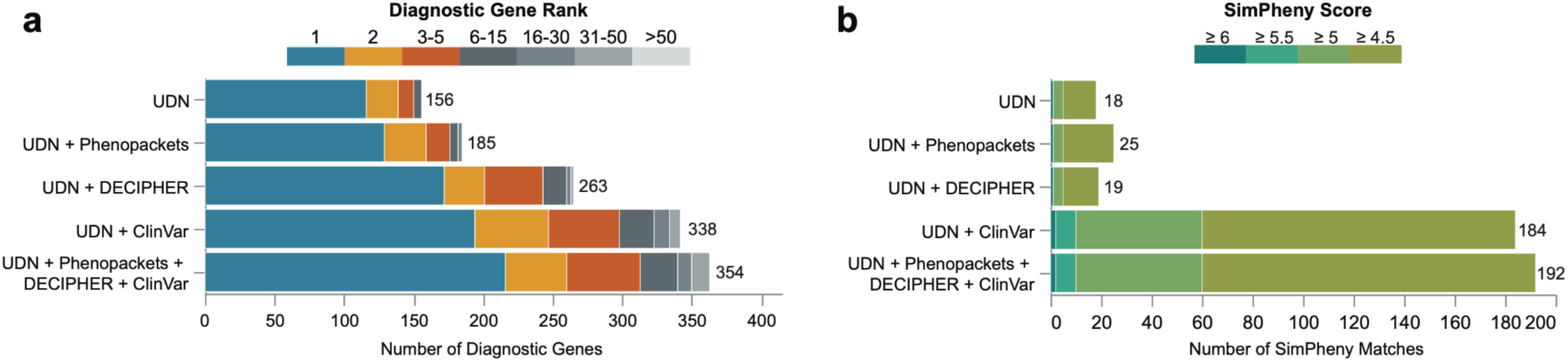
Expanding the diagnosed reference cohort increases recovery of diagnostic genes and high-confidence SimPheny candidates. **a)** SimPheny performance ranking known diagnostic genes as the diagnosed reference cohort expands. Numbers indicate the total number of diagnostic gene instances recovered (out of 415) for each cohort. **b)** Number of high-confidence SimPheny matches (*SimPheny Score* ≥ 4.5) identified in the discovery cohort as the reference cohort expands. Numbers reflect the total count of high-confidence matches identified for each reference dataset.

Across the benchmarking probands, the expanded reference cohort produced 98,423 patient-to-patient matches, including 7,869 TP matches involving the proband’s diagnostic gene (**Supplementary Table 5**). Because a single diagnostic gene instance could match multiple diagnosed reference cases, the number of TP matches substantially exceeds the number of recoverable diagnostic gene instances. Despite the larger and more heterogeneous reference cohort, the FDR at the high-confidence threshold (*SimPheny Score* ≥ 4.5) remained comparable: 16.3% versus 17.7% using the UDN-only reference cohort (**Supplementary Table 5; Table 1**; **Supplementary Fig. 11**). However, expanding the reference cohort also shifted many TP matches into lower score ranges, reducing the proportion exceeding the high-confidence threshold from 18.8% to 5.0%.

We next applied SimPheny to the 1,447 undiagnosed probands in our discovery cohort using the expanded reference cohort, and identified 309,834 patient-to-patient candidate matches (**Supplementary Table 5**). Of the 17 probands with high-confidence (SimPheny Score ≥ 4.5) candidate matches identified using the UDN-only reference cohort, 13 gained additional matches in the same gene following reference expansion, including five of the seven subsequently confirmed diagnoses (**Supplementary Table 6**). Overall, reference expansion increased the number of high-confidence candidate matches in the discovery cohort from 18 to 192 (**Fig. 6b**; **Supplementary Fig. 12**), substantially expanding the pool of candidates available for downstream clinical review.

### An interactive implementation enables real-time patient-to-patient matching

We developed an open-source web application implementing the SimPheny framework, *SimPheny.iobio*, available at http://simpheny.iobio.io. Users can provide HPO terms and, optionally, a candidate gene list representing an undiagnosed patient to query against selected diagnosed reference cohorts, including the UDN, ClinVar, DECIPHER, the Phenopacket Store, and Orphanet-derived disease models. When a candidate gene list is provided, SimPheny.iobio computes SimPheny Scores for matching reference patients and prioritizes candidate genes. When no candidate gene list is provided, the application supports phenotype-only searches using PhenoSim_Jaccard_ to identify phenotypically similar diagnosed patients (**Supplementary Fig. 13**). This interactive application enables researchers and clinicians to evaluate the patient-to-patient matching framework and generate gene matches with their own undiagnosed patient data.

## Discussion

We introduce a diagnostic framework that prioritizes candidate genes through direct patient-to-patient phenotypic matching. Existing phenotype-aware methods depend on curated disease models and gene-phenotype associations, which are indispensable but summarize heterogeneous clinical presentations into disease-level representations and can lag behind emerging gene-disease relationships. SimPheny instead treats diagnosed patients as empirical reference models, allowing individual clinical presentations to contribute directly to the interpretation of candidate genes in unsolved cases. By shifting the unit of diagnostic evidence from disease-level models to individual diagnosed patients, SimPheny enables emerging clinical evidence to inform diagnosis before the corresponding gene-disease relationships are fully represented in curated knowledge bases.

Our benchmarking results demonstrate that diagnosed patient-derived evidence provides substantial diagnostic value. SimPheny consistently reduced broad candidate gene lists to a small number of clinically interpretable candidate diagnoses while outperforming existing prioritization approaches. This advantage was greatest for genes with limited or absent phenotype associations, where association-dependent methods frequently ranked diagnostic genes poorly or could not evaluate them at all.

Retrospective benchmarking cohorts are inherently enriched for disorders with well-established gene-disease associations, as these cases are the most likely to have already received a molecular diagnosis. Consequently, our benchmarking likely underestimates the relative value of patient-to-patient matching for emerging, incompletely characterized, or atypically presenting disorders, where curated knowledge is sparse and patient-derived evidence may exist before formal gene-phenotype associations are established. SimPheny’s strong performance on association-limited genes is consistent with this expectation and suggests its greatest clinical utility may lie in the diagnostic settings least amenable to association-based prioritization approaches.

The translational potential of patient-to-patient matching is illustrated by its application to previously unsolved UDN cases. SimPheny identified one diagnosis which the UDN subsequently credited to SimPheny, and independently prioritized several candidates that were contemporaneously diagnosed through UDN clinical evaluation. The *RPL13* case demonstrates how patient-derived evidence can support diagnostic reanalysis as new gene-disease knowledge evolves, whereas the *KIF5B* case illustrates how matching to previously diagnosed patients can identify newly recognized disease genes before curated resources fully capture those associations.

These clinical examples also highlight an important property of the framework: SimPheny can only leverage evidence from previously diagnosed patients that are represented in the reference cohort. The diagnostic reach of patient-to-patient matching is therefore fundamentally determined by the breadth of the diagnosed reference cohort. Although expanding the reference cohort increased the number of candidate matches requiring review, it also increased the number of diagnosable genes and provided additional independent patient matches that strengthened evidence for candidate genes. As reference cohorts continue to grow, each newly diagnosed patient can serve as a reusable source of diagnostic evidence for subsequent unsolved cases, increasing the likelihood of identifying additional patients with the same disorder.

Several limitations should be considered. SimPheny prioritizes genes present in an input candidate gene list and therefore depends on the sensitivity of upstream sequencing, variant calling, and candidate gene list generation. In this study, benchmarking relied on candidate gene lists generated using an Exomiser-based workflow on protein-coding SNVs and indels. Consequently, genes absent from those lists could not be prioritized by SimPheny. This reflects a limitation of the present benchmarking strategy rather than the SimPheny framework itself, which is compatible with candidate genes generated using alternative variant filtering or prioritization approaches. Extending the benchmarking framework to structural and noncoding variants will require approaches that generate appropriate candidate variants and gene sets in these genomic contexts. The quality and completeness of phenotype annotations also influence performance. The UDN’s deep phenotyping provides a strong evaluation cohort but may not fully reflect the quality of phenotyping in routine clinical practice. Nevertheless, SimPheny maintained strong performance across heterogeneous external reference datasets, suggesting robustness to variability in phenotype curation and supporting its utility beyond the UDN.

Together, these findings establish patient-to-patient matching as a scalable framework for phenotype-driven rare disease diagnostics. As diagnosed reference cohorts continue to expand, patient-derived evidence is poised to become an increasingly powerful component of rare disease diagnostics and disease gene discovery. Extending this framework to undiagnosed-to-undiagnosed patient matching may further accelerate the characterization of novel Mendelian disorders and the discovery of previously unrecognized gene-phenotype relationships.

## Online Methods

### Overview of the analytical workflow

Our study followed a multistep workflow to integrate genomic and phenotypic data from the Undiagnosed Diseases Network (UDN) and evaluate the performance of SimPheny (**Fig. 1**). First, we harmonized exome and genome sequencing data across the UDN cohort to ensure consistent variant representation. We then assembled and curated UDN participant phenotype data, removing uninformative features to improve downstream similarity analyses. Candidate gene lists were then generated using Exomiser and, together with patient phenotypes, used as inputs to SimPheny. SimPheny computes patient-to-patient phenotypic similarity and identifies overlapping candidate genes with diagnosed reference cases, assigning each match a statistical score. We benchmarked SimPheny using a cohort of diagnosed UDN probands to establish confidence thresholds and then applied this framework to undiagnosed probands to identify candidate diagnostic genes.

### Harmonization of exome and genome sequencing data

Cohort-level WGS and WES variant datasets from UDN participants were generated to provide consistent variant representation across all cases. For WGS, unaligned, paired-end FASTQ files from 5,412 unique samples corresponding to 5,353 individuals from 1,772 families enrolled in the UDN as of November 2023 were aligned to human reference hg38 (with decoys and all alt contigs) and processed to produce per-sample GVCF files in the Amazon Web Services (AWS) cloud using the Clinical Genome Analysis Pipeline (CGAP, https://cgap.hms.harvard.edu/). Per-sample GVCFs were then egressed to the local Harvard institutional cluster, where single nucleotide variants (SNVs) and short insertions/deletions (indels) were jointly called across all samples using Sentieon. For WES, unaligned FASTQ pairs from 1,454 unique samples corresponding to 1,439 individuals from 314 families enrolled in the UDN as of June 2024 were processed using the same workflow. All processing steps were performed in parallel on the Harvard institutional cluster [39]. From these cohort-level variant datasets, we extracted multisample VCF files [40] for each UDN case, including affected probands and relevant affected and unaffected relatives.

### Phenotype curation and UDN data assembly

#### UDN cohort assembly and diagnostic categorization

We identified 2,698 UDN participants comprising probands and affected family members with HPO-encoded phenotype lists recorded during their clinical evaluations in reports retrieved from the internal UDN database on January 10, 2025. Each UDN diagnosis is assigned one of four certainty levels: *certain, highly likely, tentative,* or *low.* A *certain* diagnosis represents a confident genetic finding with minimal uncertainty, while a *highly likely* diagnosis indicates some uncertainty but is considered sufficient for clinical decision-making [28]. For cases with multigenic diagnoses, each gene-condition pair is assigned its own certainty level.

For this study, participants were considered **diagnosed** if they had *certain* or *highly likely* genetic diagnoses, and **undiagnosed** if they had no diagnosis, *tentative* or *low* certainty diagnoses, or clinical-only (i.e., lacking causal genetic variants) diagnoses. We only retained diagnoses with single gene findings (i.e., excluded diagnoses in large structural variants encompassing multiple genes). Based on this criterion, 767 participants were classified as diagnosed (731 probands and 36 relatives), and 1,931 were classified as undiagnosed. Relatives were retained in the diagnosed dataset if their phenotype sets did not exactly match those of their related proband. The 767 diagnosed participants, representing 592 unique genes, comprised our **UDN reference cohort** (**Supplementary Fig. 1**). 462 genes were implicated in a single participant, and 130 in more than one.

#### Phenotype curation

Phenotype annotations for UDN participants, Orphanet disease descriptions, and external reference datasets (ClinVar, the Phenopacket Store, and DECIPHER; **Supplementary Methods**) were curated to exclude terms related to prenatal and perinatal history. These terms were commonly pre-populated in earlier UDN data collection interfaces, resulting in their frequent inclusion in participant records, regardless of clinical relevance. Prenatal and perinatal history HPO terms do not usually describe the phenotypes of the genetic disorder under investigation. As such, these terms contribute noise to our phenotype-first analysis and so were removed from all participant and disease descriptions in this study. A complete list of removed terms is provided in the supplementary materials.

#### Disease category assignment

We defined 23 disease categories corresponding to the first-level child terms of *Phenotypic abnormality* in the HPO. This “top-level” of the HPO includes broad terms such as *Abnormality of the nervous system* and *Abnormality of the eye*, which serve as parent categories for more specific HPO terms. Due to the hierarchical structure of the HPO, any more specific term must trace to one or more of these top-level terms as an ancestor. To assign each patient to a single disease category, we initialized a score of 0 for each of the 23 top-level terms. For an HPO term set describing a patient or disease, each HPO term was mapped to its top-level ancestors, and the term’s information content (IC, derived from OMIM frequencies) was added to the score of each corresponding top-level ancestor. This approach weights rarer, more informative terms and therefore their top-level ancestor more heavily than common, nonspecific terms. The top-level term with the highest cumulative IC-based score was then assigned as the term set’s primary disease category.

### Candidate gene list generation with Exomiser

#### Input files and filtering

Standard Exomiser inputs include a multisample family VCF file [40], a corresponding pedigree file in PED format, and proband phenotype annotations in HPO format [41]. For each UDN family, multisample VCFs including probands and relatives were extracted from the harmonized WGS or WES variant datasets. VCFs were filtered to exclude variants with an alternate allele of “*”, genotype quality (GQ) < 20, or heterozygous calls with a variant allele frequency (VAF) less than 0.15 or greater than 0.85 [42, 43]. Proband phenotype terms were obtained from the UDN internal database, with prenatal and perinatal HPO terms removed as described above. Pedigree files were obtained from the cohort-level WGS or WES datasets.

#### Optimized versus default parameterization

We previously identified optimized parameter settings for running Exomiser on the UDN cohort [32]. To generate candidate gene lists for SimPheny, we ran Exomiser v14.0.0 with dataset version 2406, using the optimized parameters (an example YML file is provided on GitHub). Briefly, optimization included restricting the hiPHIVE phenotype prioritization algorithm [44] to human-only associations and incorporating REVEL [45], MVP [46], AlphaMissense [47], and SpliceAI [48] as variant pathogenicity predictors. The maximum allele frequency filter was set to 0.02 to accommodate compound heterozygous diagnoses, which often involve genetic variants with higher allele frequencies.

For benchmarking analyses, we also ran Exomiser using default settings, under which hiPHIVE integrates human, mouse, zebrafish, and protein-protein interaction (PPI) network data, and variant pathogenicity is evaluated using only REVEL and MVP.

Exomiser calculates gene-level variant and phenotype scores, which are combined to generate ranked lists of protein-coding candidate genes under multiple modes of inheritance [49]. For downstream SimPheny analyses, we **retained only genes with either a variant *or* phenotype score greater than 0.5, ensuring that variants with strong pathogenicity predictions but weak phenotypic evidence were not excluded**. No additional filtering (i.e., based on Exomiser-derived p-values or combined scores) was applied.

Critically, SimPheny’s performance depends on the candidate gene list it receives – it cannot prioritize an absent gene. Thus, the effectiveness of phenotype-first matching relies on the quality and breadth of upstream variant filtering. To account for this dependency, we deliberately retained not only top-scoring Exomiser candidates, but also genes harboring variants with strong pathogenicity predictions that were otherwise deprioritized due to limited or absent phenotypic evidence. This approach ensured that SimPheny could evaluate genes with strong variant evidence despite limited or absent phenotype-based support.

### Curation of benchmarking and discovery cohorts

#### Curation of the SimPheny benchmarking cohort

We restricted the benchmarking cohort to probands, as candidate gene lists were constructed using all available family-level variant data. Of the 767 diagnosed UDN participants in the UDN reference cohort, 557 were probands with variant data in the harmonized WES or WGS variant datasets, representing 579 diagnostic gene instances (19 probands had more than one genetic diagnosis). For probands with both WES and WGS data (n=26), WGS results were used.

Exomiser was run on each proband’s family-level VCF using curated HPO term lists and available pedigrees under optimized parameters. Genes with either a variant *or* phenotype score greater than 0.5 were retained, and the remaining list of ranked genes was assessed for presence or absence of the proband’s diagnostic gene(s), regardless of the specific variant or inheritance model contributing to that gene’s score. Of the 579 diagnostic gene instances, 415 (71.7%) were prioritized at any rank. The remaining 164 diagnostic gene instances were not prioritized, primarily due to dataset or algorithmic limitations: 33% (n=54) involved structural variants, which are not represented in the jointly called datasets, 30% (n=50) involved noncoding variants, which are excluded by Exomiser’s protein-coding focus, and 22% (n=36) were missed due to pedigree inaccuracies (e.g., relatives misclassified as unaffected despite mild phenotypes) (**Supplementary Fig. 6**). Probands whose diagnostic genes were absent from the Exomiser output were excluded, as SimPheny cannot recover genes not included in input candidate lists. We note that this exclusion reflects a limitation of the input data rather than of SimPheny itself.

After these exclusions, the final **SimPheny benchmarking cohort** comprised 404 probands with 415 diagnostic gene instances (nine multi-diagnosis probands remained; **Supplementary Fig. 1**; **Supplementary Fig. 14a**). Each proband was assigned an unranked candidate gene list using the filtered Exomiser results to use as input for SimPheny, mirroring the candidate gene sets available for undiagnosed patients in real diagnostic workflows (**Fig. 1**). Candidate gene lists contained a median of 103 genes before filtering, and 41 after applying the 0.5 variant/phenotype gene score requirement (**Supplementary Fig. 7a**).

#### Curation of the SimPheny discovery cohort

To evaluate SimPheny in a translational context, we assembled a discovery cohort of undiagnosed UDN probands with available variant data. Of the 1,931 undiagnosed participants, 1,447 probands had WGS or WES data present in the harmonized, jointly called datasets across various family structures (**Supplementary Fig. 1; Supplementary Fig. 14b**). For probands with both WGS and WES data (n=108), we used the WGS results.

Exomiser was run on each proband’s family-level VCF with curated HPO phenotype lists using optimized parameters. Each proband’s candidate list contained a median of 119 unique genes prior to filtering. After retaining only genes with either a variant or phenotype score greater than 0.5, the candidate lists contained a median of 42 genes (**Supplementary Fig. 7b**). These filtered lists were used as candidate gene lists to run SimPheny.

### SimPheny framework

#### Phenotypic similarity metric (PhenoSim_Jaccard_)

To quantify the similarity between HPO term sets, we developed PhenoSim_Jaccard_, a modified Jaccard index that incorporates the information content (IC) of HPO terms.

For two HPO terms *T_1_* and *T_2_*, their phenotypic similarity is defined as the intersection of their ancestral terms divided by the union of their ancestral terms:

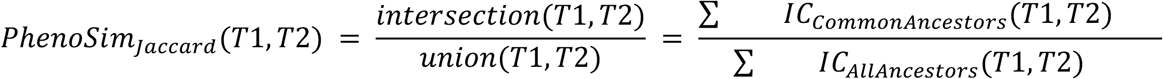

The intersection is defined as the sum of IC values for all ancestors shared by *T_1_* and *T_2_* (including the terms themselves), while the union is the sum of IC values across all ancestors of both terms. Scores range from 0 to 1, with 1 representing exact matches. Unlike the classic Jaccard index, which considers only set overlap, PhenoSim_Jaccard_ rewards shared ancestry between terms in the ontology, particularly when those ancestors are highly specific, represented by high IC values.

The IC of a term *T* is defined as:

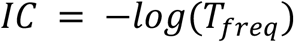

Where *T_freq_* is the proportion of OMIM diseases annotated with the term. IC values were computed using the pyHPO python package [50], and term-to-term similarities were aggregated into set-to-set similarities using the *funSimAvg* method implemented in this package [29]. Briefly, *funSimAvg* computes the similarity between two HPO term sets by averaging, for each term in one set, the highest semantic similarity it achieves with any term in the other set.

#### Definition of SimPheny Matches

For each query proband, SimPheny identifies matches as overlaps between the proband’s Exomiser-derived candidate gene list and the diagnostic genes of reference patients, supported by phenotypic similarity between the two individuals. Each such overlap defines a **SimPheny Match** (**Fig. 1**). Self-matches were excluded when the UDN cohort was used as the reference dataset to avoid trivial rediscovery by a proband matching with themself.

A single gene could yield multiple matches if several reference cases are diagnosed in that gene. Each match is scored independently based on the phenotypic similarity to the corresponding reference case, and these gene-level matches were later aggregated into SimPheny Gene Scores for downstream ranking (see below). Conversely, diagnostic genes present in only a single UDN individual cannot be validated in benchmarking with the UDN reference cohort, as no independent reference case exists for comparison.

### Calculation of the SimPheny Score

To quantify the significance of each SimPheny match, we calculated two empirical p-values: (1) *pheno p*, the probability that the observed phenotypic similarity score (PhenoSim_Jaccard_) between a query and reference case could occur by chance, and (2) *gene p*, the probability that the matched gene would appear in the query’s Exomiser-derived candidate list by chance based on cohort-wide expectations. These two measures were then combined into the *SimPheny Score*, which jointly captures phenotypic concordance and gene-level specificity.

#### Phenotypic similarity significance (*pheno p*)

To generate a null distribution for phenotypic similarity, we performed 10,000 Monte Carlo simulations per query-reference SimPheny match. In each simulation, a randomized phenotype list was generated for the query proband by sampling without replacement from the full corpus of HPO annotations in the reference dataset (42,575 annotations across 5,149 unique terms in the UDN). Sampling was weighted by term frequency to preserve population-level biases. Each simulated list was capped at 10 terms to reduce computational burden, but matched the original list length if shorter. Each simulated term list was then compared to the reference’s observed term list using the PhenoSim_Jaccard_ metric.

The *pheno p* was defined as:

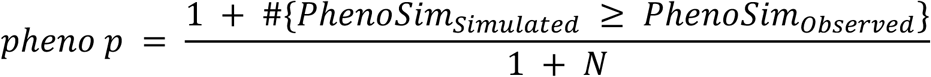

Where N = 10,000 simulations. The +1 correction prevents zero-valued p-values.

Although Monte Carlo sampling assumes independence between HPO terms, the hierarchical structure of the HPO introduces correlations. As a result, some simulated lists may partially capture the query patient’s true phenotypic presentation, potentially skewing p-values despite incorrect or overly general terms.

#### Gene match specificity (*gene p*)

To account for genes that are frequently prioritized by Exomiser – such as large genes or those associated with many nonspecific phenotypes – we generated an empirical null distribution to model the expected frequency with which each gene appears in candidate gene lists.

We compiled a corpus of all genes returned by Exomiser across 1,851 UDN probands (404 diagnosed and 1,447 undiagnosed), retaining duplicate gene entries to preserve the empirical frequency distribution. For probands with both WES and WGS data, only WGS results were used. For each query-to-reference match, we simulated 10,000 gene lists by sampling with replacement from this corpus. The number of genes in each simulated list matched the length of the proband’s filtered Exomiser-derived candidate gene list (genes with variant or phenotype score greater than 0.5).

The *gene p* was defined as:

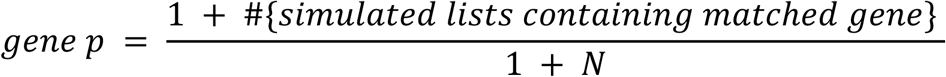

Where N = 10,000 simulations. The presence or absence of the gene was recorded for each simulated list, regardless of how many times it appeared in the sampled list. Genes can be returned by Exomiser multiple times (e.g., different modes of inheritance), but for simplicity and interpretability, we reduced this to a binary presence/absence.

This sampling approach reflects the natural frequency distribution of genes prioritized by Exomiser across the UDN cohort. Genes frequently prioritized appear more often in the corpus and thus are more likely to appear in the simulated lists. This strategy provides an empirically calibrated null model that adjusts for biases inherent to the tool, including effects of gene size, annotation density, and other Exomiser-specific biases.

#### Unifying match significance using Empirical Brown’s Method

To generate a unified measure of statistical support for each SimPheny match, we combined the *pheno p* and *gene p* into a single **combined p-value** using Empirical Brown’s Method (EBM) [51]. EBM extends Fisher’s method [52] for combining p-values by incorporating the empirical covariance structure of the input p-values, allowing for accurate p-value combination even when the input statistics are not independent.

This adjustment is critical, as *pheno p* and gene *p* are expected to be correlated: higher phenotypic similarity (*pheno p*) may increase the likelihood that a diagnostic gene appears in the Exomiser candidate gene list (*gene p*), as similar or matching phenotypes have overlapping gene-phenotype associations used by Exomiser to score and rank genes.

Unlike Fisher’s method, which assumes independence, and Stouffer’s method [53], which requires known z-scores and correlation coefficients, EBM uses empirical estimates of correlation derived from the data and outputs a single interpretable p-value per SimPheny match. The ***combined p*** resulting from EBM represents the joint probability of observing a SimPheny match under the null model, accounting for the dependency between ***pheno p*** and ***gene p***.

To parameterize EBM, we used SimPheny matches generated from the benchmarking cohort of 404 diagnosed UDN probands against each of four reference datasets: UDN, ClinVar, the Phenopacket Store, and DECIPHER. For each reference dataset, we transformed *pheno p* and *gene p* values into z-scores using the inverse normal transformation, estimated the empirical correlation (ρ) between them, and derived dataset-specific scaling factors and degrees of freedom. These fixed parameters (**Supplementary Table 7**) were then used to calculate combined p-values for all subsequent analyses of undiagnosed probands using SimPheny queried against the appropriate reference dataset.

Each reference dataset required its own fixed EBM parameters, as the distributional properties and dependencies between *pheno p* and *gene p* vary with the structure and content of the reference cohort. For example, the UDN cohort is enriched for neurological phenotypes, and participants are more deeply phenotyped compared to ClinVar pseudopatients. Using dataset-specific calibrations ensures that SimPheny Scores are appropriately scaled and comparable across different analysis contexts. Applying a single set of EBM parameters across heterogeneous datasets would distort the null distribution and compromise the interpretability of match significance.

#### SimPheny Score definition and calibration

To provide an interpretable and generalizable measure of match significance, we defined the **SimPheny Score** as the negative log_10_ of the EBM combined p-value:

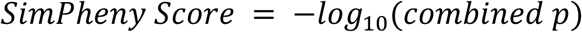

This score increases with stronger joint evidence of phenotypic similarity and gene-level specificity. A high SimPheny Score reflects a rare combination of both a phenotypically similar match and a gene that is unlikely to appear by chance in the candidate gene list.

Although multiple-testing-adjusted p-values using the Benjamini-Hochberg false discovery rate (FDR) correction [54] are also reported for completeness, these values are not suitable for generalization across cohorts. Adjusted p-values depend on the number of comparisons in a given dataset and are therefore cohort-specific. In contrast, once EBM parameters are calibrated for a reference dataset, the resulting SimPheny Score provides a portable metric that can be applied to new query patients without cohort-specific multiple-testing adjustment.

To define meaningful confidence thresholds for the SimPheny Score, we benchmarked performance across a range of scores using the diagnosed UDN benchmarking cohort. These benchmarks were used to classify matches into high-, medium-, and low-confidence categories, as described in the Results.

#### Ranking candidate genes using the SimPheny Gene Score

A single query proband can generate multiple SimPheny matches to the same gene via different reference patients. To aggregate these matches into a gene-level score for candidate gene ranking, we defined the **SimPheny Gene Score** as the average of the two highest SimPheny Scores observed for matches in that gene. This approach preserves evidence from multiple supporting matches while preventing numerous low-confidence matches from disproportionately affecting the gene score. If only a single match was observed for a given gene, the SimPheny Gene Score was set equal to that SimPheny Score.

Each proband’s candidate genes were then ranked in descending order by their SimPheny Gene Scores, producing a prioritized gene list. This ranked list was used for comparative evaluation with other prioritization tools. SimPheny Gene Scores thus provide a scalable and interpretable framework for gene prioritization that integrates multiple layers of patient similarity evidence.

### Benchmarking design

The curated benchmarking cohort of 404 diagnosed UDN probands was used as a truth set to evaluate SimPheny’s ability to recover known diagnostic genes. For each proband, we ran SimPheny on the Exomiser-derived candidate gene lists (filtered for variant or phenotype score greater than 0.5) and the proband’s HPO-encoded clinical features, querying against a reference dataset of diagnosed individuals (**Fig. 1**). Each match was classified as a true positive (TP) if the candidate gene corresponded to the proband’s known diagnostic gene, or a false positive (FP) otherwise. A SimPheny score was computed for each match as described above.

We assessed performance across a range of SimPheny Score thresholds. At each threshold (e.g., all matches with SimPheny Score ≥ 1), we calculated the following metrics:

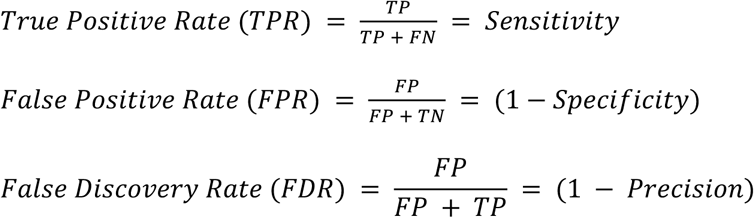

Where:

TP = number of true positive matches with scores ≥ threshold

FP = number of false positive matches with score ≥ threshold

FN = number of true positive matches with scores < threshold

TN = number of false positive matches with scores < threshold

Using these values, we generated receiver operating characteristic (ROC) curves and calculated the area under the curve (auROC) to summarize overall classification accuracy. However, ROC curves and their associated FPRs can be misleading in imbalanced datasets, where a large number of true negatives deflates the FPR even when the absolute number of false positives is high. In contrast, the FDR measures the proportion of predicted positives that are actually false at a given threshold, providing a more informative measure of performance in an unbalanced dataset. For this reason, we focused on **FDR-TPR curves** across SimPheny Score thresholds to benchmark the behavior and accuracy of the SimPheny algorithm.

### Software and computational environment

The pyHPO package [50] for ontology management was used for phenotypic similarity calculations, with a custom scoring method implemented. Exomiser version 14.0.0, with dataset version 2406, was used in the relevant analyses. All VCF subsetting and variant filtering were done with BCFtools version 1.16 [55]. AI-MARRVEL was downloaded from https://github.com/LiuzLab/AI_MARRVEL, and Phrank from https://bitbucket.org/bejerano/phrank/src/master/. Full implementation details are provided in the Supplementary Methods. SimPheny was implemented in Python and is available at https://github.com/icooperstein/SimPheny along with all code used for analysis and figure generation in this manuscript.

## Code Availability

SimPheny.iobio is freely available as an open-source web application at https://simpheny.iobio.io/, with source code hosted at https://github.com/iobio/SimPheny.iobio. SimPheny can also be run locally via https://github.com/icooperstein/SimPheny. All code used to perform the analyses, generate figures, and support the findings of this manuscript is available in the same repository.

## Acknowledgments

This study utilizes data generated by the DECIPHER community. A complete list of centers that contributed to the generation of the data is available from https://deciphergenomics.org/about/stats and via email from. DECIPHER is hosted by EMBL-EBI and funding for the DECIPHER project was provided by the Wellcome Trust [grant number WT223718/Z/21/Z]. We thank the participants and families of the Undiagnosed Diseases Network for their generous contributions and willingness to share sensitive clinical information in support of the broader research community. Those who carried out the original analysis and collection of the data bear no responsibility for the further analysis or interpretation of the data.

## Author Contributions

I.C., A.W., and G.M. contributed to the conception and design of the study, including methods and interpretation of the results. I.C. fetched metadata associated with participants. I.C. curated cohorts. S.K. performed realignment and joint calling of UDN GS and ES data. I.C. performed tool development, computational analyses, and generated all figures. E.B. developed the SimPheny.iobio web application. I.C. drafted the original manuscript. A.W., S.K., E.B., B.M., R.S., V.S., and G.T.M. contributed to manuscript review and editing. All authors reviewed the final manuscript.

## Data Availability

All deidentified exomic and genomic sequencing data, including SNV and indel variant calls, as well as corresponding phenotype data in the form of pedigree files and HPO terms, are regularly deposited in dbGaP (accession phs001232.v7.p3). Rare SNV and indel variants, as well as HPO terms, for UDN participants with genomic sequencing are also queryable in a public-facing browser: https://dbmi-bgm.github.io/udn-browser/. Variant-level data, clinical significance, and supporting evidence, as well as demographic information and phenotype information for all diagnostic variants in the UDN, are periodically submitted to ClinVar. Other relevant, patient-specific clinical information may be shared on a case-by-case basis at the discretion of the clinical team managing the case. Requests for such data can be directed to with a response expected within six weeks.

