## Supplementary Materials for "Patient-to-patient phenotype matching enables rare disease gene prioritization beyond curated gene–phenotype associations"

### Supplementary Figures

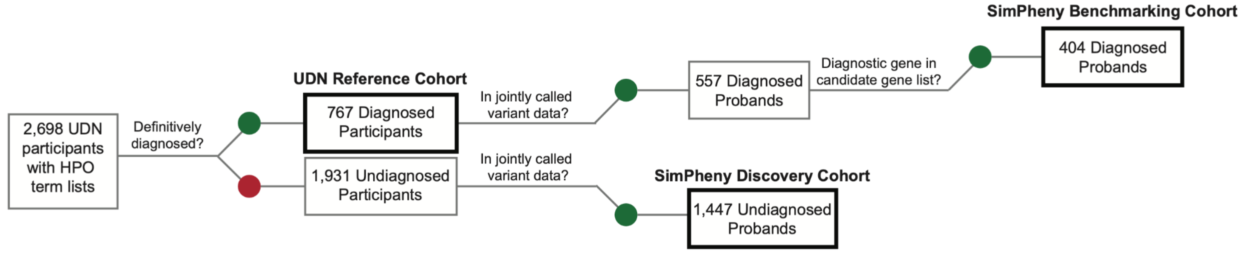


**Supplementary Figure 1: Derivation of UDN reference, discovery, and benchmarking cohorts used in SimPheny analyses.**
We extracted 2,698 UDN participants with annotated HPO terms from the internal UDN database on January 10, 2025. Of these, 767 participants had a *certain* or *highly likely* genetic diagnosis and were used as the **UDN Reference Cohort** of diagnosed participants. The remaining 1,931 were labeled as undiagnosed, including those with no genetic diagnosis, tentative or low confidence genetic findings, or clinical-only diagnoses. Among the undiagnosed participants, 1,447 probands had available family-level VCF data from jointly called WES or WGS datasets, enabling Exomiser runs to generate candidate gene lists. These probands comprised the **SimPheny Discovery Cohort**. Among the 767 diagnosed participants, 557 probands also had family-level VCF data from jointly called WES or WGS datasets. To define the **SimPheny Benchmarking Cohort**, we further filtered to retain only those whose known diagnostic gene was present in their Exomiser-derived candidate gene list, representing cases in which the diagnostic gene was recoverable from the input candidate gene list.

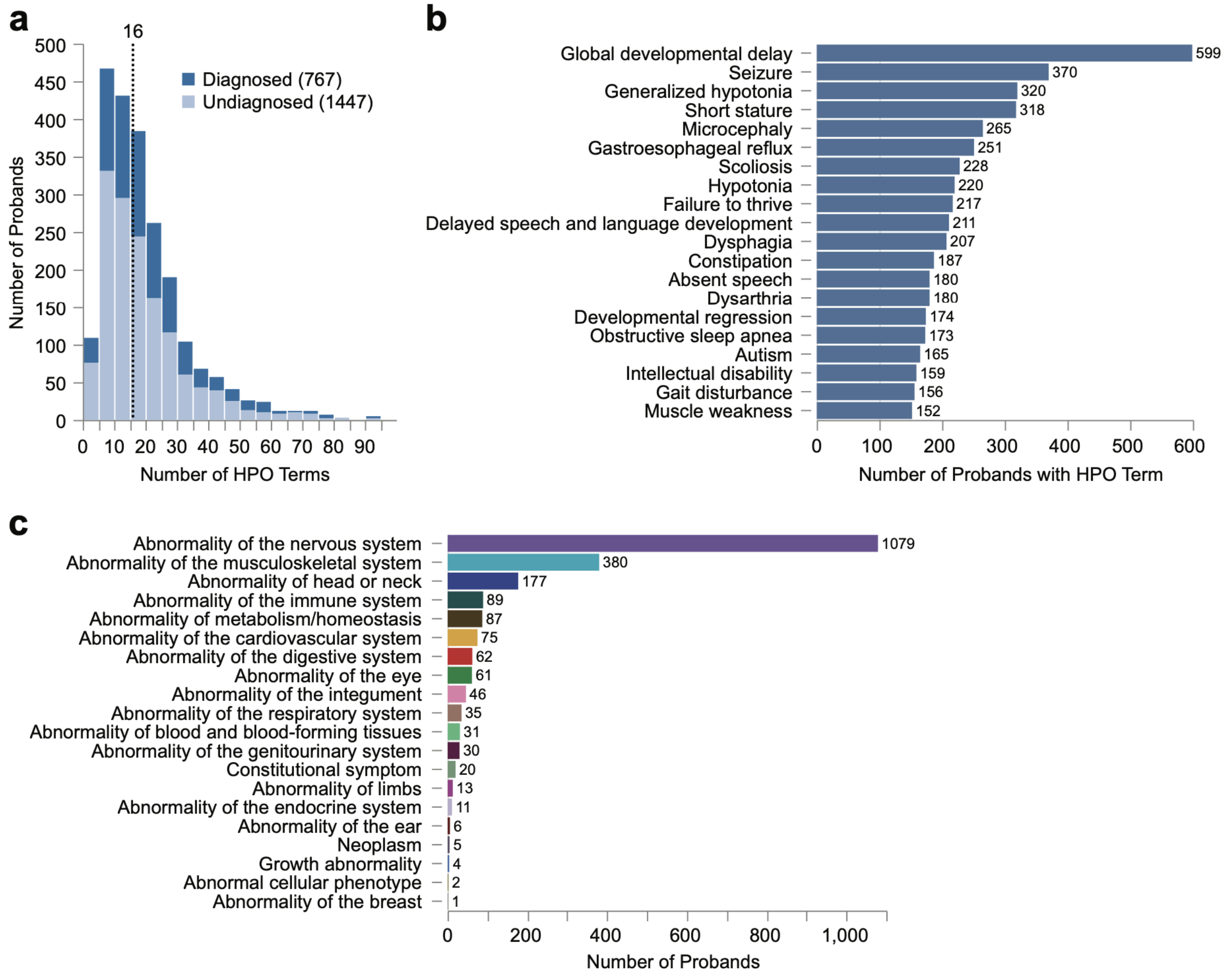


**Supplementary Figure 2: Summary of phenotype annotations in 2,214 UDN probands with jointly called variant data.**

**a)** Distribution of the number of HPO terms per proband following phenotype curation (removal of prenatal and perinatal terms). Dashed line indicates the median number of terms across all probands. Color corresponds to diagnostic status.

**b)** The 20 most frequently annotated HPO terms across the UDN cohort.

**c)** Distribution of top-level HPO disease categories assigned to probands in the UDN cohort.

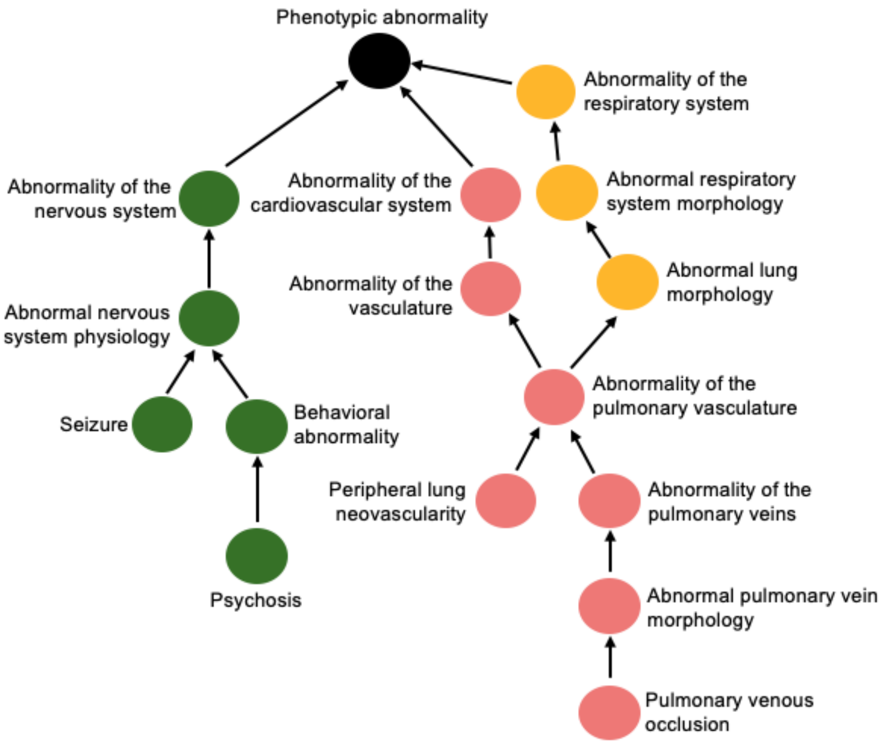


**Supplementary Figure 3: The Human Phenotype Ontology (HPO) is organized as a directed acyclic graph.**

More specific phenotype terms (child nodes) are connected to broader parent terms through directed edges toward a common root phenotype term ("Phenotypic abnormality"). Example neurological, pulmonary, and respiratory phenotype hierarchies are shown to illustrate how related phenotypes share ancestral ontology structure.

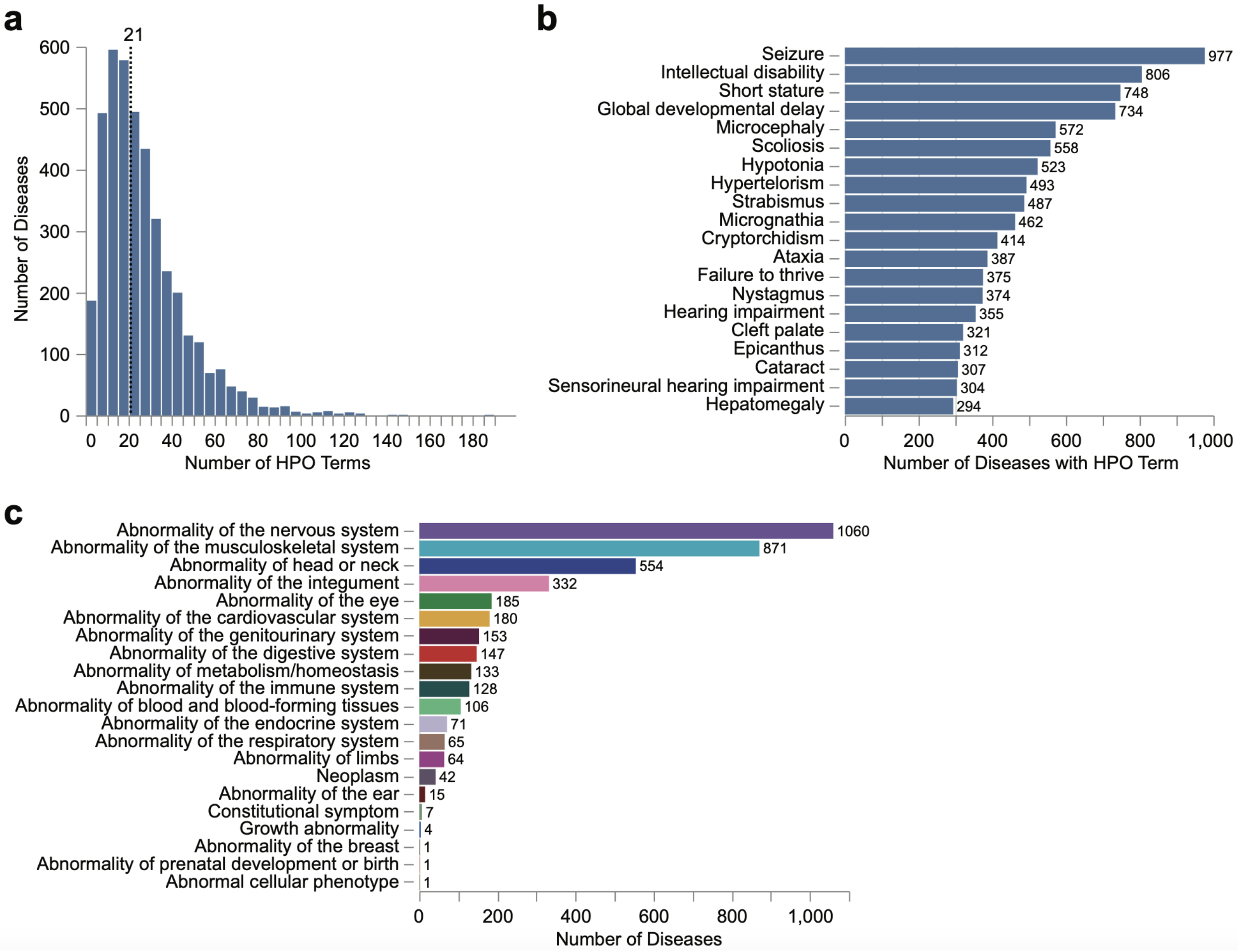


**Supplementary Figure 4: Summary of phenotype annotations in 4,120 Orphanet disease descriptions**.

**a)** Distribution of the number of HPO terms per disease following phenotype curation (removal of prenatal and perinatal terms). Dashed line indicates the median number of terms across all diseases.

**b)** The 20 most frequently annotated HPO terms across the Orphanet disease cohort.

**c)** Distribution of top-level HPO disease categories assigned to Orphanet diseases.

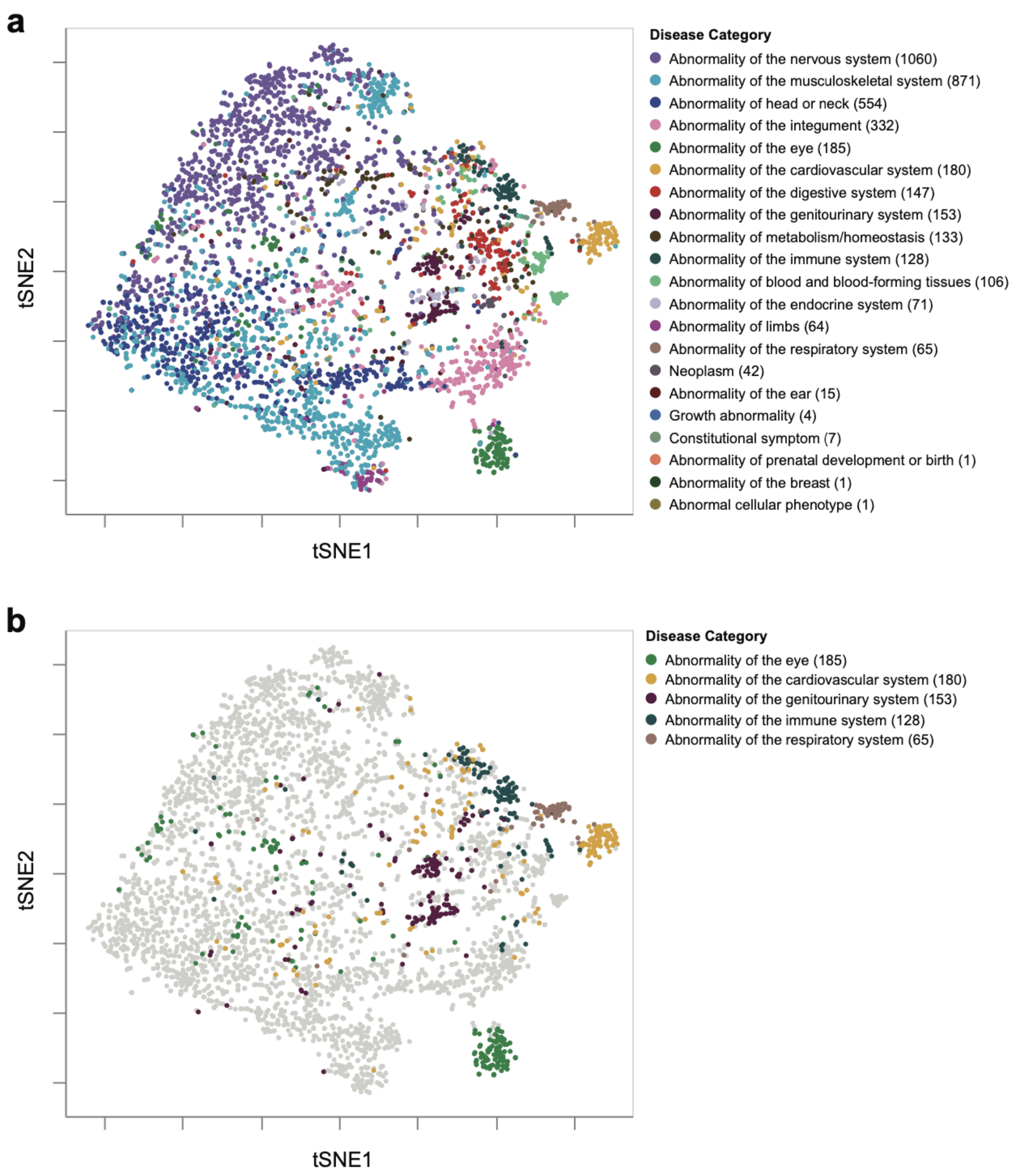


**Supplementary Figure 5: PhenoSim$_{Jaccard}$ separates rare diseases by primary system involvement.**

**a)** t-SNE projection of pairwise PhenoSim$_{Jaccard}$ scores among 4,120 Orphanet disease descriptions. Each point represents a single disease, colored by its top-level HPO category - one of 23 top-level terms directly beneath *Phenotypic abnormality* in the HPO hierarchy. These categories reflect broad physiological systems.

**b)** Same as (**a**), but only five of the 23 HPO system categories are colored to highlight distinct groups of phenotypically similar diseases affecting the same physiological system. All other diseases are shown in grey.

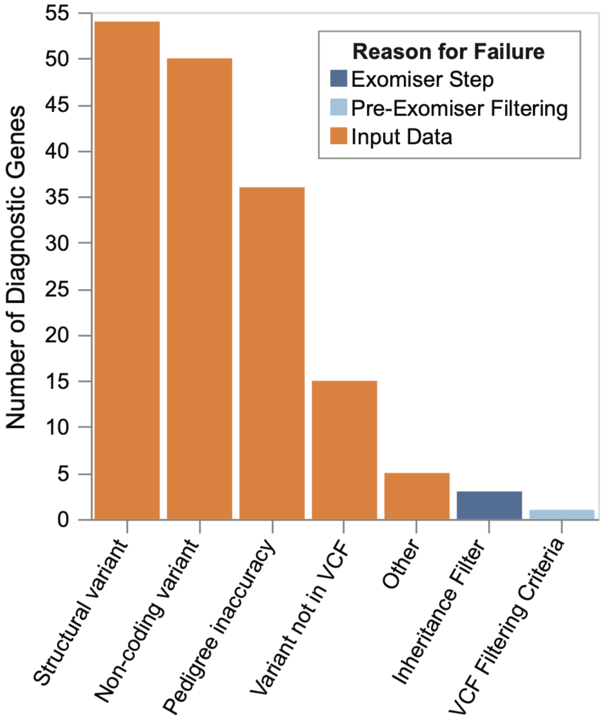


**Supplementary Figure 6: Diagnostic gene instances excluded from SimPheny benchmarking cohort.**

A total of 164 diagnostic gene instances were not prioritized when Exomiser was run on filtered family-level VCFs for 557 diagnosed UDN probands. These VCFs contain only SNV and indel variants; WGS VCFs may therefore include noncoding SNVs and indels, but Exomiser performs only protein-coding variant analysis, so noncoding variants are not found in its outputs.

**Orange**: Gene prioritization failure due to limitations in input data. These include structural or noncoding variants, or cases where the known diagnostic SNV/indel variant is not in the supplied VCF. *Other* includes complex diagnoses (i.e., multiple gene interactions) and primary diagnostic methods that are not standard sequencing (WES/WGS) tests. *Pedigree inaccuracy* refers to instances where the incorrect affected status of family members disrupted Exomiser's inheritance-based filtering. For example, in some cases, a proband inherited the causal variant maternally, but the mother was annotated as unaffected, causing the variant to be excluded.

**Dark blue**: Genes not prioritized due to a filtering or scoring step within the Exomiser algorithm itself.

**Light blue**: Genes lost during pre-Exomiser VCF filtering (ALT!=*; GQ ≥ 20; 0.15 ≤ VAF ≤ 0.85; see Methods).

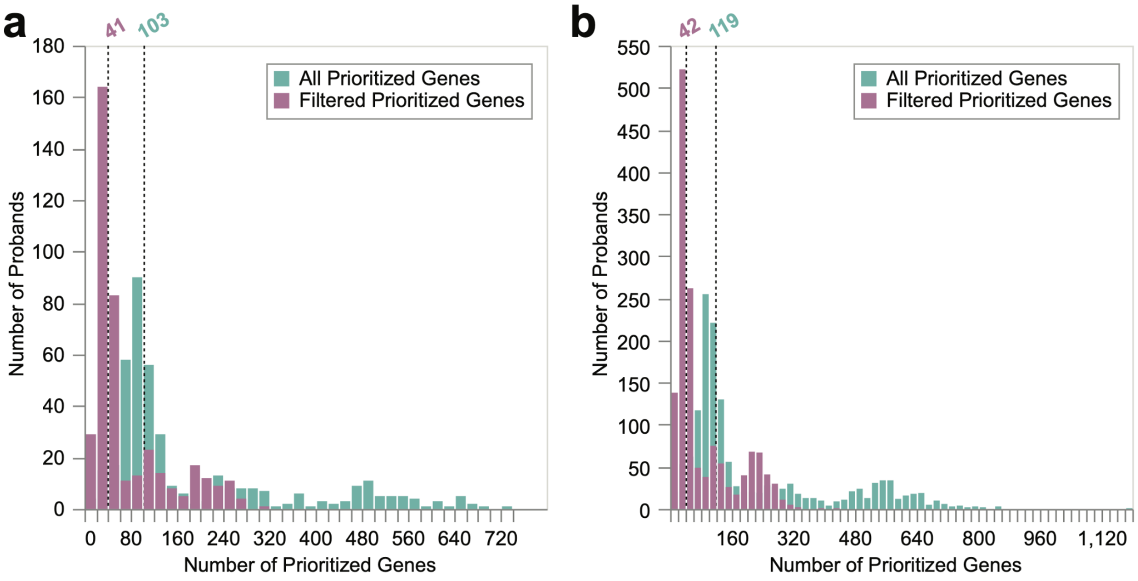


**Supplementary Figure 7: Distribution of Exomiser-prioritized gene list lengths across diagnosed and undiagnosed probands.**

Distribution of the number of unique candidate genes prioritized by Exomiser in **a)** 404 diagnosed probands in the SimPheny benchmarking cohort and **b)** 1,447 undiagnosed probands in the SimPheny discovery cohort. Green bars represent the full set of unique genes ranked by Exomiser, while purple bars reflect the number of genes remaining after applying a filter requiring either a gene-level variant score *or* phenotype score > 0.5. Dashed lines indicate the median candidate gene list length for each condition, with corresponding numeric labels.

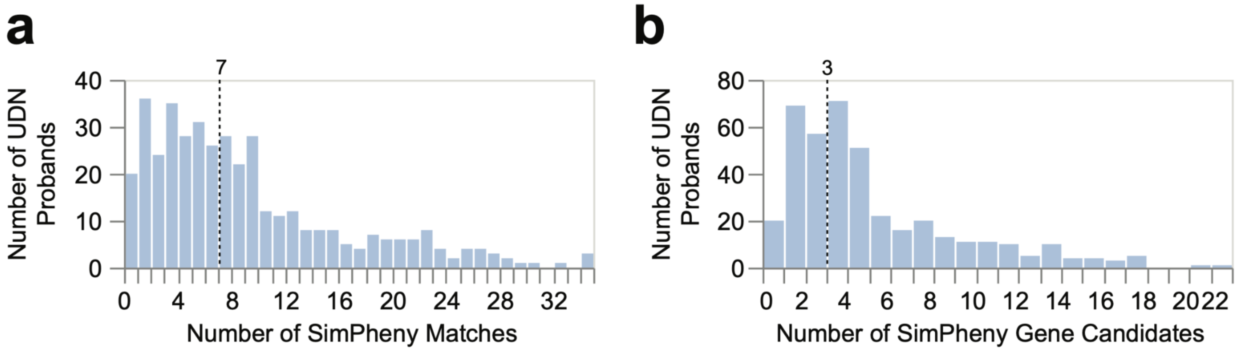


**Supplementary Figure 8: SimPheny matches and candidate genes identified against the UDN diagnosed reference dataset.**

**a)** Number of SimPheny matches identified when comparing each of the 404 diagnosed UDN probands in the benchmarking cohort against the 767 diagnosed UDN reference individuals. A single proband may have multiple SimPheny matches to the same gene if multiple reference individuals are diagnosed in that gene. Dashed line indicates the median number of SimPheny matches per proband.

**b)** Number of unique SimPheny gene candidates (i.e., distinct diagnostic genes from matched reference individuals) returned per proband. Dashed line indicates the median number of unique candidate genes per proband.

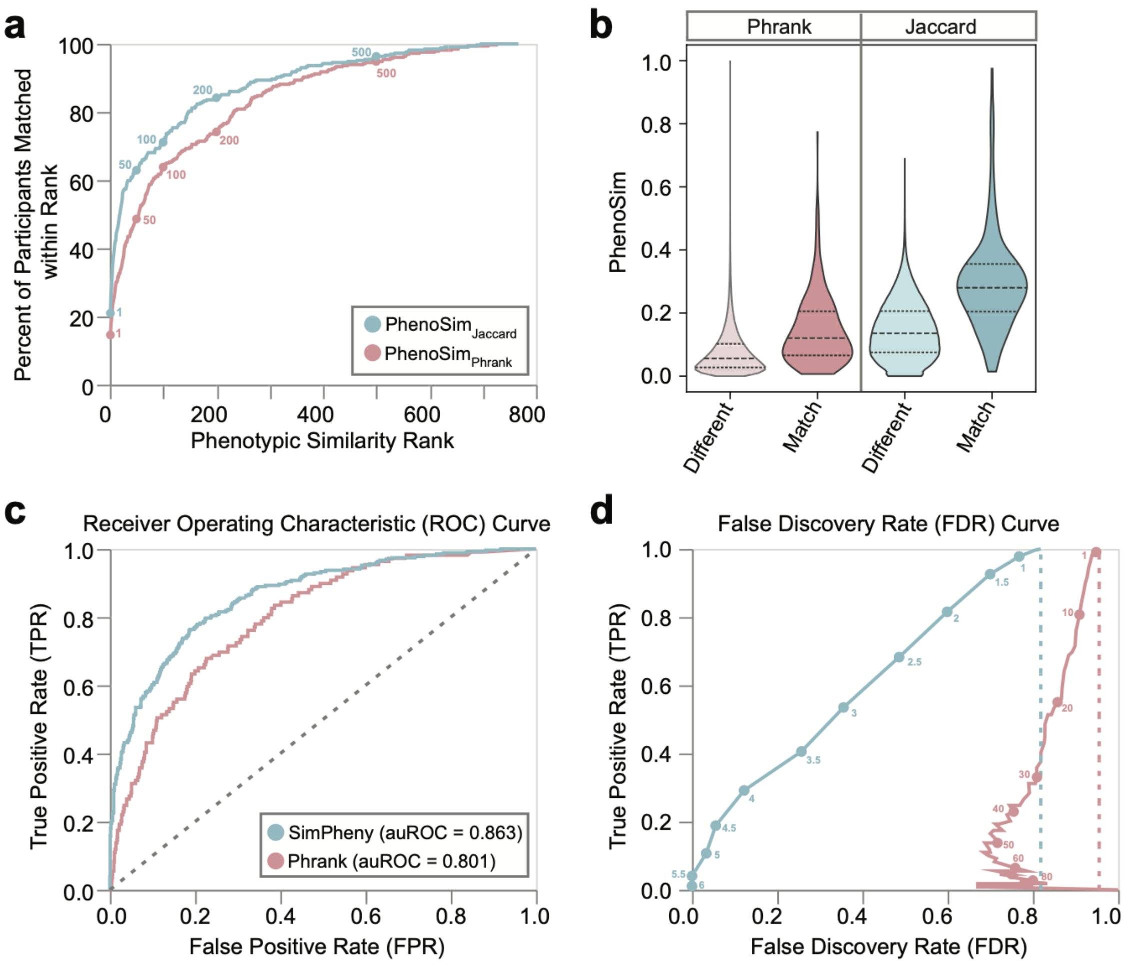


**Supplementary Figure 9: Benchmarking PhenoSim$_{Jaccard}$ against Phrank**

Pairwise phenotypic similarity scores were calculated between all 767 diagnosed UDN participants in the UDN reference cohort using either the PhenoSim$_{Jaccard}$ metric (**blue**) or the PhenoSim$_{Phrank}$ metric (**pink**).
**a**) For each participant, all others were ranked from most to least phenotypically similar (rank 1 to 766). 329 of these participants shared a diagnostic gene with at least one other participant in the cohort. We evaluated the rank at which each participant first matched to another participant with the same diagnostic gene and calculated the cumulative percentage of 329 participants whose diagnostic match was ranked at or below each rank threshold (x-axis). Rank thresholds 1, 50, 100, 200, and 500 are labeled.
**b)** Pairwise phenotypic similarity scores between all 767 diagnosed UDN participants were stratified by whether each pair shared the same diagnostic gene (“match”) or not (“different”). Results are shown separately for PhenoSim$_{Phrank}$ (left) and PhenoSim$_{Jaccard}$ (right). Dashed lines denote the 25th, 50th, and 75th percentiles of each distribution.
**c)** Receiver Operating Characteristic (ROC) curve evaluating the ability of SimPheny Scores (**blue**) and Phrank gene scores (**pink**) to distinguish true diagnostic matches from false positives. Analysis includes the subset of 156 benchmarking probands for whom SimPheny successfully recovered the diagnostic gene using the UDN reference dataset.
**d)** False Discovery Rate (FDR) curves generated by using SimPheny Score (**blue**) and Phrank gene scores (**pink**) as thresholds (labeled) for predicting true matches. Dashed lines indicate the random expectation based on the class balance in this set of 156 probands - 82.1% of SimPheny matches and 95.6% of Phrank matches are expected to be false positives under random chance.

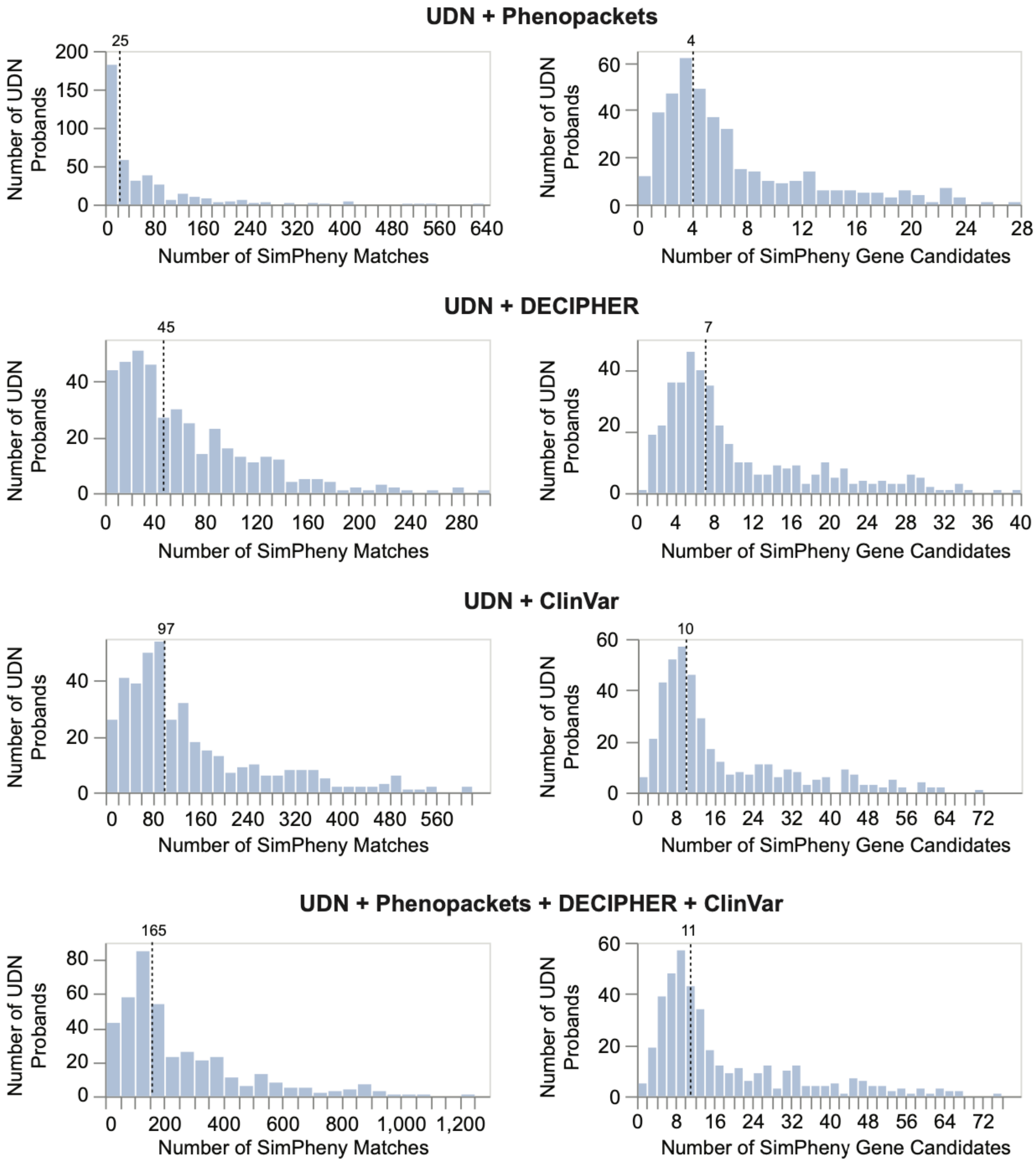


**Supplementary Figure 10: Expanding reference datasets increases SimPheny matches and candidate genes.**

**Left panel:** Number of SimPheny matches identified when comparing each of the 404 diagnosed UDN probands in the benchmarking cohort against each reference dataset in the title. A single proband may have multiple SimPheny matches to the same gene if multiple reference individuals are diagnosed in that gene. Dashed line indicates the median number of SimPheny matches per proband.

**Right panel:** Number of unique SimPheny gene candidates (i.e., distinct diagnostic genes from matched reference individuals) returned per proband. Dashed line indicates the median number of unique candidate genes per proband.

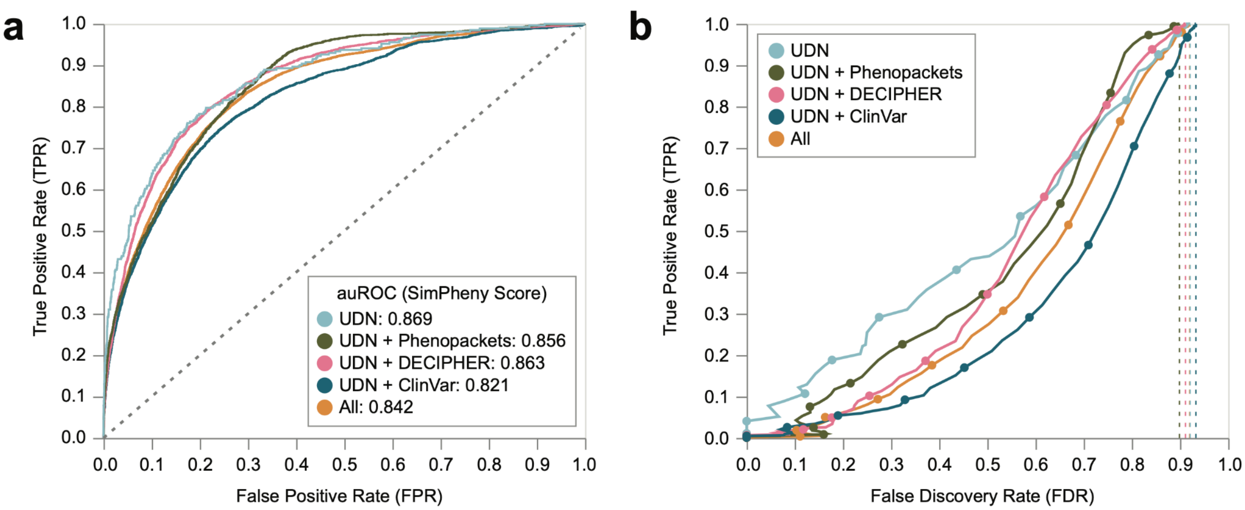


**Supplementary Figure 11: Benchmarking SimPheny Scores using expanded reference datasets.**

**a)** Receiver Operating Characteristic (ROC) curves evaluating the ability of SimPheny Scores to distinguish true diagnostic matches from false positives across different reference cohorts, using the full set of 404 UDN benchmarking probands. Curve colors correspond to the reference dataset used.

**b)** False Discovery Rate (FDR) curves assessing SimPheny Score thresholds for predicting true matches, stratified by reference cohort. Dashed lines indicate the expected FDR based on the proportion of false positives within each reference set. Curve colors match those in panel **(a)**.

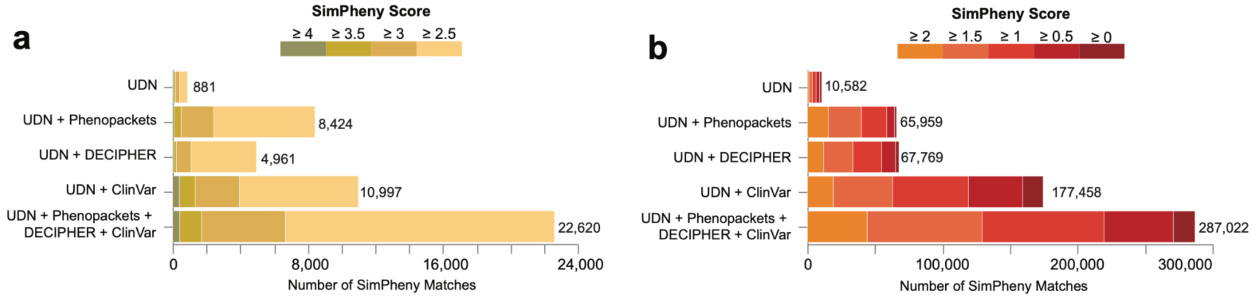


**Supplementary Figure 12: Expanding the diagnosed reference set increases medium- and low-confidence SimPheny matches**

**a)** Number of medium confidence SimPheny matches (4.5 > SimPheny Score ≥ 2.5) returned in the discovery cohort as the reference cohort expands. Numbers reflect the total count of medium confidence matches identified for each reference dataset.

**b)** Number of low confidence SimPheny matches (SimPheny Score < 2.5) returned in the discovery cohort as the reference cohort expands. Numbers reflect the total count of low-confidence matches identified for each reference dataset.

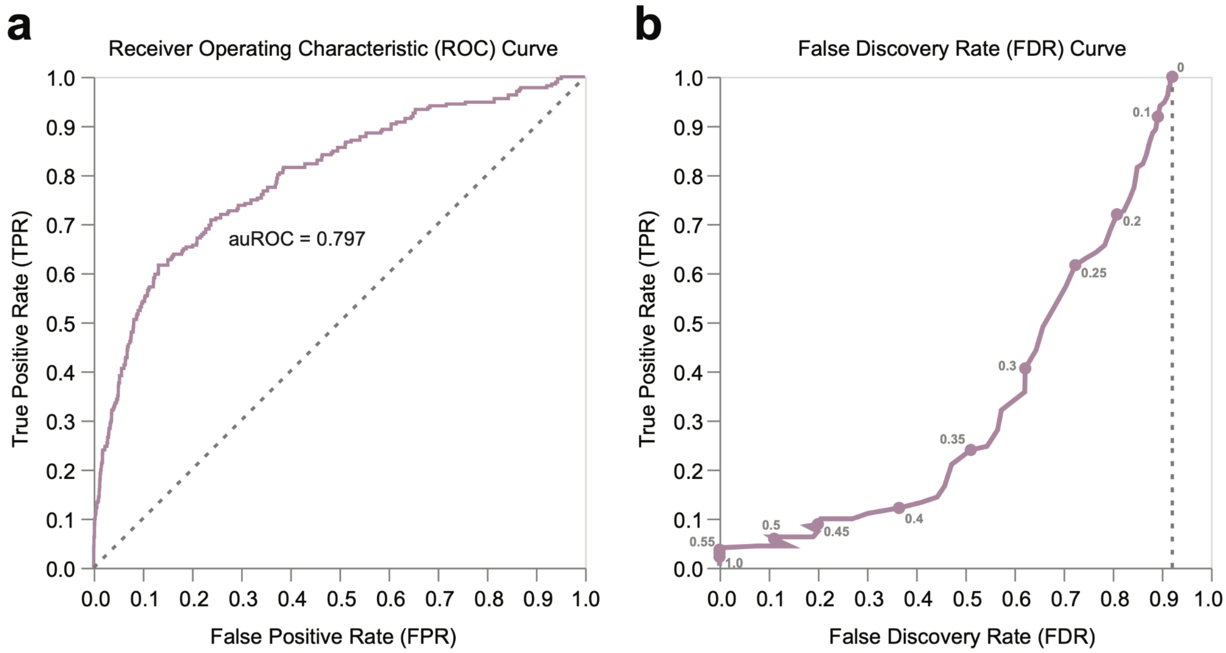


**Supplementary Figure 13: Benchmarking phenotype-only patient matching using $\mathbf{PhenoSim_{Jaccard}}$ score thresholds.**

When no candidate gene list is provided, SimPheny.iobio cannot compute SimPheny Scores because no gene-level overlap can be evaluated. In this setting, phenotype-only searches rely on $PhenoSim_{Jaccard}$ scores to rank phenotypically similar diagnosed reference patients. We therefore benchmarked the ability of $PhenoSim_{Jaccard}$ scores to distinguish true positive patient pairs sharing the same genetic diagnosis from false positive pairs with different diagnoses across varying score thresholds.

**a)** Receiver Operating Characteristic (ROC) curve evaluating the performance of the $PhenoSim_{Jaccard}$ metric across varying thresholds.

**b)** False Discovery Rate (FDR) curve. Each point corresponds to a $PhenoSim_{Jaccard}$ threshold (in increments of 0.05) ranging from 0 (top right) to 1 (bottom left). The dashed line indicates the random expectation, where 92.1% of matches are expected to be false positives due to class balance.

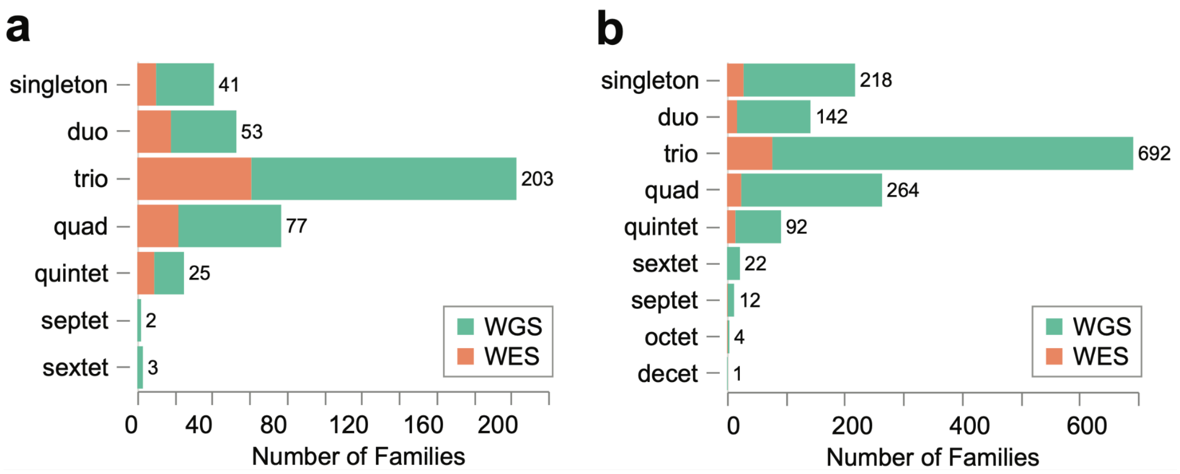


**Supplementary Figure 14: Breakdown of sequencing data and family structures used in SimPheny analyses.**

**a)** Family structures* of 404 diagnosed UDN probands used in the SimPheny benchmarking cohort. Colors indicate whether Exomiser was run on jointly called WES data (orange) or WGS data (green).

**b)** Family structures* of 1,447 undiagnosed UDN probands used in the SimPheny discovery cohort, similarly colored by WES data (orange) or WGS (green) data.

*Family structure refers to the number of closely related individuals included in the analysis. For example, “trio” indicates any family structure consisting of three related family members, not necessarily a parent–parent–child trio.

### Supplementary Tables

**Supplementary Table 1: Summary statistics for SimPheny in the benchmarking cohort against the UDN reference dataset**

| SimPheny Score | TP Matches | FP Matches | TPR | FDR | SimPheny Matches | SimPheny Candidates | Confidence |
|---|---|---|---|---|---|---|---|
| ≥ 6.0 | 3 | 0 | 1.1% | 0.0% | 1 | 1 | High |
| ≥ 5.5 | 11 | 0 | 4.1% | 0.0% | 1 | 1 | High |
| ≥ 5.0 | 29 | 4 | 10.7% | 12.1% | 1 | 1 | High |
| ≥ 4.5 | 51 | 11 | 18.8% | 17.7% | 1 | 1 | High |
| ≥ 4.0 | 79 | 30 | 29.2% | 27.5% | 1 | 1 | Medium |
| ≥ 3.5 | 110 | 85 | 40.6% | 43.6% | 2 | 1 | Medium |
| ≥ 3.0 | 145 | 191 | 53.5% | 56.8% | 2 | 2 | Medium |
| ≥ 2.5 | 185 | 400 | 68.3% | 68.4% | 2 | 2 | Medium |
| ≥ 2.0 | 221 | 825 | 81.5% | 78.9% | 3 | 2 | Low |
| ≥ 1.5 | 251 | 1480 | 92.6% | 85.5% | 5 | 3 | Low |
| ≥ 1.0 | 265 | 2266 | 97.8% | 89.5% | 7 | 4 | Low |
| ≥ 0.5 | 270 | 2882 | 99.6% | 91.4% | 8 | 5 | Low |
| ≥ 0.0 | 271 | 3171 | 100% | 92.1% | 9 | 5 | Low |

SimPheny was run on 404 diagnosed UDN probands in the benchmarking cohort using a reference dataset of 767 diagnosed UDN participants. Each row summarizes results at a given SimPheny Score threshold (leftmost column). “SimPheny Matches” refers to the *average* number of matches per proband with at least one match at that threshold. “SimPheny Candidates” refers to the average number of unique *gene* matches per proband. True positive (TP), false positives (FP), true positive rate (TPR), and false discovery rate (FDR) were calculated based as described in **Figure 3**. Confidence tiers were derived from benchmarking performance.

**Supplementary Table 2: SimPheny candidate gene ranking performance compared to alternate tools**

| Tool | k=1 | k=2 | k=3 | k=5 | k=10 | k=15 | k=20 | k=30 | k=50 | k>50 | Not Prioritized |
|---|---|---|---|---|---|---|---|---|---|---|---|
| **SimPheny** | 116 | 139 | 148 | 150 | 154 | 156 | 156 | 156 | 156 | 0 | 0 |
| **AI-MARRVEL** | 112 | 127 | 129 | 131 | 135 | 140 | 142 | 143 | 145 | 5 | 6 |
| **Exomiser** | 116 | 131 | 137 | 140 | 146 | 148 | 153 | 154 | 156 | 0 | 0 |
| **Phrank** | 61 | 70 | 78 | 88 | 98 | 102 | 103 | 104 | 104 | 0 | 52 |
| **Exomiser (default)** | 105 | 118 | 123 | 130 | 138 | 143 | 145 | 147 | 148 | 8 | 0 |

Each tool was evaluated on its ability to prioritize the true diagnostic gene for 156 probands in the SimPheny benchmarking cohort that were successfully recovered by querying SimPheny against the UDN reference dataset. The table reports the number of cases in which the diagnostic gene was ranked within the top *k* candidates (e.g., top 1, 3, 5, 10, etc). “Not prioritized” indicates cases where the diagnostic gene was not returned by the tool. “k>50” indicates that the gene was ranked lower than 50th. “Exomiser (default)” refers to Exomiser run with default configuration, while “Exomiser” refers to our optimized parameters (see Methods).

**Supplementary Table 3: Extended match details for high-confidence SimPheny candidates (SimPheny Score ≥ 4.5)**

| Proband | Pheno Sim | Pheno Rank | Candidate Gene | Pheno *p* | Gene *p* | Combined *p* | Combined *p* (adj) | Exomiser Rank Pheno Variant | SimPheny Score | Status | ClinVar Classification |
|---|---|---|---|---|---|---|---|---|---|---|---|
| **UDN1** | **0.352** | **1** | **RPL13** | **0.0001** | **0.0002** | **8.12E-07** | **0.009** | **1<br>0.5753<br>0.9964** | **6.091** | **D** | **US** |
| **UDN1** | **0.314** | **6** | **RPL13** | **0.0006** | **0.0003** | **5.68E-06** | **0.013** | **1<br>0.5753<br>0.9964** | **5.246** | **D** | **US** |
| **UDN2** | **0.464** | **8** | **AFG3L2** | **0.0005** | **0.0004** | **6.23E-06** | **0.013** | **1<br>0.4589<br>0.9618** | **5.206** | **CD** | **CC** |
| **UDN3** | **0.316** | **4** | **KIF5B** | **0.0005** | **0.0006** | **8.90E-06** | **0.013** | **10<br>0.0000<br>0.9993** | **5.050** | **CD** | **US** |
| UDN4 | 0.417 | 7 | MED12 | 0.0001 | 0.0034 | 9.94E-06 | 0.013 | 1<br>0.6754<br>0.8488 | 5.003 | U | CC |
| UDN5 | 0.660 | 1 | SET | 0.0001 | 0.0038 | 1.10E-05 | 0.013 | 225<br>0.5883<br>0.0996 | 4.960 | U | LB |
| UDN6 | 0.326 | 111 | HCFC1 | 0.0002 | 0.0022 | 1.25E-05 | 0.013 | 9<br>0.7773<br>0.0939 | 4.904 | U | B/LB |
| UDN7 | 0.380 | 19 | WDR73 | 0.0009 | 0.0005 | 1.27E-05 | 0.013 | 3<br>0.5534<br>0.4540 | 4.896 | U | US |
| **UDN8** | **0.384** | **7** | **WARS2** | **0.0006** | **0.0010** | **1.64E-05** | **0.013** | **3<br>0.5783<br>0.6148** | **4.786** | **CD** | **CC; CC** |
| UDN9 | 0.405 | 3 | TRAF3 | 0.0001 | 0.0061 | 1.66E-05 | 0.013 | 5<br>0.4845<br>0.8908 | 4.780 | RC | N/A |
| UDN10 | 0.498 | 2 | MTOR | 0.0001 | 0.0066 | 1.78E-05 | 0.013 | 1<br>0.7921<br>0.8473 | 4.750 | U | LB |
| UDN11 | 0.506 | 5 | ANK3 | 0.0001 | 0.0067 | 1.80E-05 | 0.013 | 17<br>0.6570<br>0.0547 | 4.744 | U | B |
| UDN12 | 0.344 | 22 | MED12 | 0.0010 | 0.0007 | 1.87E-05 | 0.013 | 1<br>0.6391<br>0.6827 | 4.727 | U | CC |
| UDN13 | 0.513 | 2 | ERCC4 | 0.0001 | 0.0073 | 1.94E-05 | 0.013 | 135<br>0.0000<br>0.8000 | 4.711 | U | LB |
| **UDN14** | **0.316** | **89** | **GLUL** | **0.0030** | **0.0003** | **2.34E-05** | **0.013** | **7<br>0.0000<br>0.9944** | **4.631** | **CD** | **LP** |
| **UDN15** | **0.340** | **52** | **SPTBN1** | **0.0006** | **0.0018** | **2.74E-05** | **0.013** | **1<br>0.7130<br>0.8266** | **4.562** | **TD** | **US** |
| UDN16 | 0.301 | 22 | RFC1 | 0.0008 | 0.0014 | 2.83E-05 | 0.013 | 12<br>0.0000<br>0.8000 | 4.548 | U | N/A |
| **UDN17** | **0.262** | **8** | **UBA1** | **0.0001** | **0.0115** | **2.90E-05** | **0.013** | **2<br>0.5002<br>0.9721** | **4.538** | **CD** | **CC** |

This table expands on the high-confidence matches presented in Table 2, showing all SimPheny Matches with SimPheny Score ≥ 4.5 returned in the undiagnosed UDN discovery cohort (n=1,447 probands) queried against the UDN reference cohort (n=767 diagnosed participants). Each row represents a candidate gene match prioritized by SimPheny, including additional columns not shown in Table 2: the empirical *pheno p* and *gene p* values, as well as *combined p* and adjusted *p-values (*via Empirical Brown's Method), Exomiser rank and variant/phenotype scores (from optimized parameter run), and PhenoSim$_{Jaccard}$ similarity metrics. Manual review classifications are reported in the "Status" column: D = Diagnostic; CD = Contemporaneously Diagnosed; RC = Rejected Candidate; TD = Tentative Diagnosis (excluded from diagnosed cohort due to uncertainty at time of curation); U = Unresolved. Bolded rows indicate the seven candidate genes considered diagnostic upon clinical review. US = Uncertain Significance; CC = Conflicting classifications of pathogenicity; LB = Likely benign; B/LB = Benign/Likely benign; B = Benign; LP = Likely pathogenic; N/A = not in ClinVar

**Supplementary Table 4: Impact of reference dataset composition on recovery of diagnostic genes and SimPheny match characteristics**

| Reference Cohort (Size) | Unique Genes Represented in Cohort | Diagnostic Gene Instances Recovered | Diagnostic Gene Instances in Top 5 Candidates | Mean True Matches | Udx Probands with at Least 1 Match | Mean SimPheny Matches | Mean SimPheny Candidates |
|---|---|---|---|---|---|---|---|
| **UDN (767)** | 592 | 156 (37.6%) | 150 (96.2%) | 2 | 1,250 (86.4%) | 9 | 5 |
| **UDN + Phenopacket Store (7,218)** | 939 | 185 (44.6%) | 176 (95.1%) | 13 | 1,318 (91.1%) | 58 | 7 |
| **UDN + DECIPHER (5,343)** | 1,335 | 263 (63.4%) | 243 (92.4%) | 8 | 1,378 (95.2%) | 55 | 9 |
| **UDN + ClinVar (12,564)** | 2,676 | 338 (81.4%) | 298 (88.2%) | 12 | 1,408 (97.3%) | 134 | 16 |
| **UDN + Phenopacket Store + DECIPHER + ClinVar (23,591)** | 2,939 | 354 (85.3%) | 313 (88.4%) | 23 | 1,410 (97.4%) | 225 | 17 |

SimPheny was run on the benchmarking cohort of 404 diagnosed UDN probands using five different reference cohorts: UDN-only (n=767), or UDN combined with diagnosed cases from the Phenopacket Store, DECIPHER, ClinVar, or all three. For each reference cohort, we report: (i) the number of unique genes represented by the reference cohort patients, (ii) the number and proportion of diagnostic gene instances recovered out of 415 (SimPheny match identified at any score), (iii) the number of diagnostic gene instances ranked among the top five gene candidates for each proband based on SimPheny Gene Scores and subsequent gene ranking, and (iv) the mean number of SimPheny matches per recovered diagnostic gene instance. For the discovery cohort of 1,447 undiagnosed probands, we report the number of probands with at least one match as well as the mean number of SimPheny matches and the mean number of SimPheny gene candidates per proband.

**Supplementary Table 5: SimPheny matches summary statistics across SimPheny Score thresholds in benchmarking and discovery cohorts using the full expanded reference dataset.**

| SimPheny Score | Undiagnosed Matches | TP Matches | FP Matches | TPR | FDR | SimPheny matches | SimPheny Candidates | Confidence |
|---|---|---|---|---|---|---|---|---|
| ≥ 6.0 | 2 | 5 | 0 | 0.06% | 0% | 1 | 1 | High |
| ≥ 5.5 | 10 | 32 | 4 | 0.4% | 11.1% | 2 | 1 | High |
| ≥ 5.0 | 60 | 147 | 17 | 1.9% | 10.4% | 2 | 1 | High |
| **≥ 4.5** | **192** | **395** | **77** | **5.0%** | **16.3%** | **3** | **1** | High |
| ≥ 4.0 | 602 | 737 | 276 | 9.4% | 27.2% | 3 | 1 | Medium |
| ≥ 3.5 | 1905 | 1382 | 865 | 17.6% | 38.5% | 4 | 2 | Medium |
| ≥ 3.0 | 6857 | 2416 | 2758 | 30.7% | 53.3% | 8 | 3 | Medium |
| ≥ 2.5 | 22812 | 4046 | 8150 | 51.4% | 66.8% | 21 | 5 | Medium |
| ≥ 2.0 | 66949 | 6014 | 20826 | 76.4% | 77.6% | 53 | 8 | Low |
| ≥ 1.5 | 152217 | 7255 | 44021 | 92.2% | 85.9% | 112 | 13 | Low |
| ≥ 1.0 | 242539 | 7711 | 72052 | 98.0% | 90.3% | 178 | 16 | Low |
| ≥ 0.5 | 293741 | 7838 | 86545 | 99.6% | 91.7% | 214 | 17 | Low |
| ≥ 0.0 | 309834 | 7869 | 90554 | 100% | 92.0% | 225 | 17 | Low |

SimPheny was run on 1,851 UDN probands - 404 diagnosed individuals in the benchmarking cohort and 1,447 undiagnosed individuals in the discovery cohort - using the full extended reference dataset combining 767 diagnosed UDN participants, 6,451 Phenopacket Store cases, 4,576 DECIPHER cases, and 11,797 ClinVar pseudopatients. Each row summarizes results at a given SimPheny Score threshold (leftmost columns). True positive (TP) and false positive (FP) matches were determined from the benchmarking cohort, while “Undiagnosed Matches” refers to all matches returned from the discovery cohort at or above each threshold. TPR (true positive rate) and FDR (false discovery rate) were calculated using benchmarking data only. “SimPheny Matches” and “SimPheny Candidates” indicate the average number of matches or candidate genes per proband (of 1,851) from among those with at least one match at each score threshold. Confidence tiers (high, medium, low) were assigned based on benchmarking performance.

**Supplementary Table 6: SimPheny candidate matches for 17 probands selected for diagnostic review**

| Proband | Gene | Non-UDN Matches | Non-UDN Match SimPheny Scores | UDN Matches | UDN Match SimPheny Scores |
|---|---|---|---|---|---|
| **UDN1** | **RPL13** | **1** | **6.26** | **3** | **6.09, 5.25, 4.45** |
| **UDN2** | **AFG3L2** | **4** | **3.31, 3.17, 2.93, 2.62** | **2** | **5.21, 2.68** |
| **UDN3** | **KIF5B** | **0** | **-** | **1** | **5.05** |
| UDN4 | MED12 | 27 | 3.91, 3.53, 3.2, 3.17, 2.92, 2.85, 2.8, 2.73, 2.69, 2.62, 2.62, 2.61, 2.59, 2.59, 2.56, 2.56, 2.5, 2.48, 2.46, 2.45, 2.4, 2.39, 2.35, 2.28, 2.26, 2.21, 2.17 | 1 | 5.0 |
| UDN5 | SET | 4 | 2.73, 2.72, 2.47, 2.32 | 2 | 4.96, 2.65 |
| UDN6 | HCFC1 | 5 | 2.71, 2.49, 2.38, 2.38, 2.38 | 1 | 4.9 |
| UDN7 | WDR73 | 3 | 4.51, 4.07, 3.13 | 1 | 4.9 |
| **UDN8** | **WARS2** | **4** | **3.29, 2.98, 2.83, 2.79** | **2** | **4.79, 2.42** |
| UDN9 | TRAF3 | 0 | - | 1 | 4.78 |
| UDN10 | MTOR | 19 | 4.01, 3.25, 3.13, 2.55, 2.5, 2.49, 2.47, 2.42, 2.37, 2.36, 2.35, 2.28, 2.27, 2.25, 2.24, 2.2, 2.19, 2.02, 1.84 | 4 | 4.75, 2.19, 1.93, 1.87 |
| UDN11 | ANK3 | 0 | - | 1 | 4.74 |
| UDN12 | MED12 | 27 | 3.18, 3.06, 3.05, 3.04, 3.02, 3.01, 2.94, 2.9, 2.9, 2.88, 2.82, 2.8, 2.77, 2.74, 2.73, 2.72, 2.66, 2.6, 2.59, 2.56, 2.55, 2.52, 2.51, 2.48, 2.4, 2.39, 2.26 | 1 | 4.73 |
| UDN13 | ERCC4 | 3 | 2.43, 2.25, 1.28 | 1 | 4.71 |
| **UDN14** | **GLUL** | **9** | **3.77, 3.76, 3.71, 3.54, 3.41, 3.33, 3.31, 3.14, 3.1** | **1** | **4.63** |
| **UDN15** | **SPTBN1** | **2** | **2.39, 2.24** | **2** | **4.56, 3.32** |
| UDN16 | RFC1 | 1 | 2.31 | 3 | 4.55, 2.37, 2.28 |
| **UDN17** | **UBA1** | **0** | **-** | **2** | **4.54, 3.82** |

This table summarizes all SimPheny matches identified for the 17 undiagnosed UDN probands' high-confidence gene candidates (SimPheny Score ≥ 4.5) prioritized in the discovery cohort for diagnostic review (**Table 2**), focusing on the breakdown of matches returned from UDN reference individuals versus those returned from non-UDN cases in the expanded reference dataset (Phenopacket Store, DECIPHER, ClinVar). For each proband, we list the candidate gene, the number of non-UDN matches and their corresponding SimPheny Scores, as well as the number and scores of UDN-based matches. Bolded rows highlight the SimPheny high-confidence candidates that were confirmed diagnostic upon review.

**Supplementary Table 7: Fixed parameters for the Empirical Brown's Method used to combine pheno and gene p-values.**

| Cohort | ρ | Scaling factor | Degrees of freedom |
|---|---|---|---|
| UDN | 0.140654089465608 | 1.070327044732804 | 3.737175491999793 |
| ClinVar | 0.06694913859450671 | 1.0334745692972533 | 3.8704387305049393 |
| Phenopacket Store | 0.16390782006306615 | 1.0819539100315332 | 3.6970151527835613 |
| DECIPHER | 0.12171835819912691 | 1.0608591790995634 | 3.7705287174826747 |

For each reference cohort, the average pairwise correlation (ρ) among input p-values was used to derive the fixed scaling factor and degrees of freedom required by Empirical Brown's Method (EBM). These fixed values were applied to all SimPheny Score calculations using that reference dataset, enabling consistent and computationally efficient estimation of combined significance scores across large-scale queries. See Methods for full description of EBM application and parameter derivation.

### Supplementary Methods

#### External reference dataset curation for SimPheny matching

Unlike the UDN consortium, where patients are deeply phenotyped and carefully curated, external reference resources vary considerably in quality, structure, and phenotype annotation standards. Below, we describe the curation and processing of external datasets used in SimPheny analyses, including the ClinVar, Phenopacket Store, and DECIPHER diagnosed reference cohorts and Orphanet disease descriptions used for related phenotypic similarity analyses.

##### ClinVar "pseudopatients" data assembly

ClinVar provides variant-level submissions (SCVs), which may include HPO-coded clinical features. Unlike true patient datasets, ClinVar does not provide identifiers linking multiple variants to the same individual. As a result, SCVs were treated as "pseudopatients," though some likely derive from the same person.

We parsed the public ClinVar XML aggregated by VCV records, which aggregates data for a specific variant across all submitted records. We identified 57,541 SCVs, represented by ClinicalAssertion elements, with HPO annotations in the April 3, 2025 release file (https://www.ncbi.nlm.nih.gov/clinvar/docs/ftp_primer/). We applied the following filter steps:

1. VariationType: *Indel or single nucleotide variant (SNV)*
2. GermlineClassifications: *Pathogenic* or *Likely pathogenic*
3. ReviewStatus: *"Criteria provided, single submitter"* (one gold star)

These criteria retained 13,768 SCVs. In an effort to remove multiple gene submissions from a single patient, we removed submissions with exactly matching metadata across the following fields:
*SubmitterName, HPOTerms, Ethnicity, AgeMin, AgeMax, Sex, GeneSymbol*.
However, this metadata is not available for every submission.

Since the UDN regularly submits solved cases to ClinVar, we removed 295 SCVs with the *SubmitterName* field "Undiagnosed Diseases Network, NIH" to avoid self-matching against the benchmarking cohort. We excluded 28 unrecognized HPO terms (likely submission errors), which led to 41 SCVs losing terms and three losing all annotations. 814 pseudopatients had prenatal/perinatal HPO terms removed, and this led to 11 pseudopatients losing all annotated terms entirely.

Following these steps, 11,797 pseudopatient SCVs remained to make up our **ClinVar reference dataset**. Because no patient identifiers exist, we cannot confirm whether some represent compound heterozygous diagnoses or multigenic submissions per individual.

Across this set:

- 2,497 unique genes were represented
- 892 genes were implicated in a single pseudopatient
- 1,605 genes were implicated in more than one pseudopatient

#### Phenopacket Store data assembly

We downloaded version 0.1.19 of the Phenopacket Store from the Monarch Initiative (https://monarch-initiative.github.io/phenopacket-store/). Each phenopacket represents a single individual. We parsed 6,668 phenopackets from this resource and applied the following inclusion criteria:

- At least one “present” HPO term (some listed only “not present” terms).
- At least one associated diagnostic gene

This left us with 6,452 individual phenopackets. Filtering prenatal and perinatal terms led to the removal of HPO terms from 368 phenopackets, with one losing all terms entirely. Our final **Phenopacket reference dataset** included 6,451 individuals.
Across this set:

- 421 unique genes were represented
- 87 genes were implicated in a single patient
- 334 genes were implicated in multiple patients

Each phenopacket was assigned a single diagnostic gene; no phenopacket had more than one.

#### DECIPHER data assembly

We downloaded the DECIPHER bulk data file on August 29, 2025, containing 14,505 SNV and indel variant-level entries with associated phenotypes. Unlike ClinVar, each variant in DECIPHER is mapped to a deidentified patient ID, enabling patient-level aggregation and filtering.

We applied the following inclusion criteria:

1. Pathogenicity: *Pathogenic or Likely pathogenic*
2. Contribution: *Full or Partial*
3. At least three HPO terms per patient

Applying these filters yielded 5,651 entries across 5,232 unique patient identifiers. Prenatal/perinatal HPO terms were removed from 447 individuals, leading three to lose all terms. The final **DECIPHER reference dataset** included 4,576 individuals. Most individuals (4,500) had diagnostic variants in only one gene, but 76 individuals had diagnostic variants in two or more genes.

Across this set:

- 999 unique genes were represented
- 421 were implicated in a single patient
- 578 genes were implicated in multiple patients

### Other data resources

These external resources were not used as reference cohorts for SimPheny matching but supported related analyses such as $PhenoSim_{Jaccard}$ validation.

#### Orphanet disease descriptions

Orphanet data was downloaded on March 30, 2024 from the Orphanet scientific knowledge base (https://www.orphadata.com/).

We combined data from three Orphadata tables:

- *Genes associated with rare diseases*
- *Clinical signs and symptoms in rare diseases*
- *Natural history of rare diseases*

Each HPO term in Orphanet is annotated with one of the following frequency categories:

- Always present (100%)
- Very frequent (80-99%)
- Frequent (30-79%)
- Occasional (5-29%)
- Rare (1-4%)
- Excluded (0%)

We removed any phenotype terms with frequency “Excluded (0%).” We also excluded any disease description labeled “Multigenic/multifactorial” or “Oligogenic” in the natural history file. After filtering, 4,120 disease descriptions remained, of which 2,344 had at least one associated gene.

### Comparative tools

#### Phrank

Phrank is a phenotype-driven prioritization method that takes two inputs: a list of patient phenotypes encoded in Human Phenotype Ontology (HPO) terms and a list of candidate genes [29]. For each gene in the list, Phrank computes a similarity score between the patient’s HPO term set and the phenotype set annotated to that gene. Each phenotype’s contribution is weighted inversely by the number of genes known to cause that phenotype, giving more weight to rarer, more specific terms. The result is a ranked list of candidate genes, with higher scores indicating stronger phenotypic concordance.

We downloaded Phrank from the official repository at https://bitbucket.org/bejerano/phrank/ and used it to calculate gene scores for each gene in the benchmarking probands’ Exomiser-derived candidate gene lists using their HPO terms as input. Gene scores produced by Phrank were used in place of SimPheny Scores to generate ROC curves and FDR-TPR curves, enabling direct comparison of performance. We also used these scores to generate Phrank-ranked candidate gene lists for each proband to benchmark against SimPheny Gene Score-based rankings.

Phrank's performance is contingent on its underlying knowledge base of gene-phenotype associations. In its default implementation, these are drawn from HPO Annotations [49], which are primarily based on OMIM. As a result, ***Phrank can only score genes that are present in this knowledge base.***

In addition to gene-based scoring, Phrank's similarity measure can also be applied directly to compare two sets of HPO terms. We leveraged this functionality to compute a $PhenoSim_{Phrank}$ score, which we used to benchmark our own phenotypic similarity measure ($PhenoSim_{Jaccard}$) against an established ontology-based similarity measure.

#### AI-MARRVEL

AI-MARRVEL (Model organism Aggregated Resources for Rare Variant Exploration) integrates genomic, phenotypic, and functional annotations to prioritize candidate variants for Mendelian disorders [21]. This tool supports analysis in singleton mode (proband-only) or trio mode (proband and both parents), but does not support other family structures. Since our benchmarking cohort includes a heterogeneous mix of family configurations (**Supplementary Fig. 14**), we ran AI-MARRVEL in singleton mode on all 404 probands to ensure consistency.

We downloaded AI-MARRVEL from the official GitHub repository at https://github.com/LiuzLab/AI_MARRVEL and ran it locally on singleton VCFs extracted from the jointly called WES and WGS datasets. Each run incorporated the proband's HPO term list as phenotypic input. AI-MARRVEL returned a ranked list of candidate variants for each proband, which we mapped to their corresponding genes to generate gene-level rankings. These were then compared to SimPheny Gene Score-based gene rankings in benchmarking analyses.

#### Perinatal/prenatal HPO terms removed

HP:0012188 | Hyperemesis gravidarum
HP:0008071 | Maternal hypertension
HP:0009800 | Maternal diabetes
HP:0030244 | Maternal fever in pregnancy
HP:0100622 | Maternal seizure
HP:0011438 | Maternal teratogenic exposure
HP:0100603 | Toxemia of pregnancy
HP:0011436 | Abnormal maternal serum screening
HP:0001511 | Intrauterine growth retardation
HP:0001562 | Oligohydramnios
HP:0001561 | Polyhydramnios
HP:0001558 | Decreased fetal movement
HP:0010519 | Increased fetal movement
HP:0001787 | Abnormal delivery
HP:0001622 | Premature birth
HP:0001518 | Small for gestational age
HP:0001520 | Large for gestational age
HP:0003561 | Birth length less than 3rd percentile
HP:0003517 | Birth length greater than 97th percentile
HP:0011451 | Primary microcephaly
HP:0004488 | Macrocephaly at birth
HP:0002643 | Neonatal respiratory distress
HP:0006579 | Prolonged neonatal jaundice
HP:0002033 | Poor suck
HP:0001998 | Neonatal hypoglycemia
HP:0040187 | Neonatal sepsis
HP:0011410 | Caesarian section
HP:0030364 | Secondary Caesarian section
HP:0030369 | Induced vaginal delivery
HP:0001788 | Premature rupture of membranes
HP:0001194 | Abnormalities of placenta or umbilical cord
